# Forecasting the COVID-19 epidemic integrating symptom search behavior: an infodemiology study

**DOI:** 10.1101/2021.03.09.21253186

**Authors:** Alessandro Rabiolo, Eugenio Alladio, Esteban Morales, Andrew I McNaught, Francesco Bandello, Abdelmonem A. Afifi, Alessandro Marchese

## Abstract

**Background:** Previous studies have suggested associations between trends of web searches and COVID-19 traditional metrics. It remains unclear whether models incorporating trends of digital searches lead to better predictions.

**Methods:** An open-access web application was developed to evaluate Google Trends and traditional COVID-19 metrics via an interactive framework based on principal components analysis (PCA) and time series modelling. The app facilitates the analysis of symptom search behavior associated with COVID-19 disease in 188 countries. In this study, we selected data of eight countries as case studies to represent all continents. PCA was used to perform data dimensionality reduction, and three different time series models (Error Trend Seasonality, Autoregressive integrated moving average, and feed-forward neural network autoregression) were used to predict COVID-19 metrics in the upcoming 14 days. The models were compared in terms of prediction ability using the root-mean-square error (RMSE) of the first principal component (PC1). Predictive ability of models generated with both Google Trends data and conventional COVID-19 metrics were compared with those fitted with conventional COVID-19 metrics only.

**Findings:** The degree of correlation and the best time-lag varied as a function of the selected country and topic searched; in general, the optimal time-lag was within 15 days. Overall, predictions of PC1 based on both searched termed and COVID-19 traditional metrics performed better than those not including Google searches (median [IQR]: 1.43 [0.74-2.36] vs. 1.78 [0.95-2.88], respectively), but the improvement in prediction varied as a function of the selected country and timeframe. The best model varied as a function of country, time range, and period of time selected. Models based on a 7-day moving average led to considerably smaller RMSE values as opposed to those calculated with raw data (median [IQR]: 0.74 [0.47-1.22] vs. 2.15 [1.55-3.89], respectively).

**Interpretation:** The inclusion of digital online searches in statistical models may improve the prediction of the COVID-19 epidemic.

**Funding:** EOSCsecretariat.eu has received funding from the European Union’s Horizon Programme call H2020-INFRAEOSC-05-2018-2019, grant Agreement number 831644.

## INTRODUCTION

COVID-19 is a new entity, and the dynamics of its propagation are difficult to predict. In the absence of compelling evidence, health and political decisions have been strongly driven by a wide variety of statistical models and simulation scenarios to forecast the COVID-19 epidemic. Still, large variations exist among the different models with respect to the predicted number of infected people, time to reach a peak of new cases, course of the epidemic, and identification of outbreaks.^1^ One key limitation of such models is that they rely heavily on the number of confirmed infected subjects, who are usually seeking medical attention due to moderate to severe symptoms. However, confirmed cases are most likely only a small proportion of the true number of cases as the vast majority of infected individuals have an asymptomatic or mildly symptomatic disease.^2^

There is increasing interest in the potential of ‘big data’ analysis to predict future areas of COVID-19 outbreaks and incidence of cases based on symptom search behaviors. In the past, search query data have been used to facilitate early detection and near real-time estimates of flu and Dengue.^3^ These seminal works were helpful in introducing later improvements to predictive models that might help guide health authorities to mount rapid responses, and to design more efficient surveillance programs for COVID-19.^4^ A few studies have shown a correlation between Google Trends of medical terms searches and COVID-19 metrics,^5^ suggesting that incorporating Google Trends data into conventional metrics could lead to better nowcasting and forecasting of the COVID-19 epidemic.

In this study, we systematically evaluate patterns of web queries for COVID-19 clinical manifestations and develop an open-access web application for exploring their correlations with COVID-19 propagation. We implement models integrating conventional COVID-19 metrics with Google Trends data and compare them to those not containing Google Trends data. The results of this study provide a framework for digital surveillance of COVID-19 using open-access big data.

## METHODS

### Data Collection

COVID-19 daily new confirmed, cumulative number, and number per million of cases and deaths for all available world countries were automatically exported from the COVID-19 Data Repository by the Center for System Science and Engineering at Johns Hopkins University (source: https://github.com/CSSEGISandData/COVID-19).^6^ The selected countries used as case studies are given in the results section below. Countries choice was arbitrary, and the following principles were adopted: representation of the five continents; inclusion of countries where the COVID-19 epidemic had different levels of severity and different evolutions over time; inclusion of countries where Google is the preferred search engine; exclusion of countries with limited access to the internet; exclusion of countries where one or more Google Trends topic had only zero or missing vales in the selected time frame; exclusion of countries whose reliability in terms of data reporting has been questioned.

As data were fully anonymized and publicly available, no ethical approval was required.

Google Trends API was used to extract trends of Google searches for the most common COVID-19 signs and symptoms in those countries.^7^ For each search term, geographic region, and time frame selected, Google Trends outputs an ‘interest over time’ (IOT) index, which estimates the relative search volume on a normalized scale from 0 (no searches) to 100 (search term popularity peak). Twenty topics were identified on the basis of the most frequent signs and symptoms of COVID-19 and included: abdominal pain, ageusia, anorexia, anosmia, bone pain, chills, conjunctivitis, cough, diarrhea, eye pain, fatigue, fever, headache, myalgia, nasal congestion, nausea, rhinorrhea, shortness of breath, sore throat, tearing.^8-11^ Google Trends queries were carried out with the «topic» function, which includes all the related terms sharing the same concept in different languages. This approach ensures that the frequency of searches for closely related symptom types are appropriately grouped together.

For each country and search term, data were automatically exported as csv files for two pre-specified timeframes: (i) five years weekly data from 12/Apr/2015 to 5/Apr/2020 to study the long-term pattern of searched term, and (ii) daily data from 22/Jan/2020 to 20/Dec/2020. As Google Trends allows daily data exportation up to 9 month, daily data were reconstructed by means of an overlapping method.^12^

### Data Analysis

Interest-over-time values for the five-year interval were used to distinguish topics with a significant deviation from their long-term pattern from the onset of COVID-19 epidemic. For seasonal queries, trends were isolated from seasonal and random components with an additive decomposition method (Supplementary Figure 1); for non-seasonal queries, trends were extracted by smoothing the time series with a one-year moving average. Decomposition plots were visually inspected, and topics with no clear change in their five-year trends from January 2020 were excluded from the subsequent analyses.

The relationship between the daily IOT values for the selected topics and COVID-19 confirmed deaths and new cases were investigated in the shorter time frame indicated above. Relationships between IOT values of each topic and the number of new daily confirmed cases or deaths per million were visually assessed with line graphs. Changes in IOT values over time were visually assessed with streamgraphs. To smooth daily fluctuations in both IOT values and number of new cases, plots were generated using a 7-day moving average.

Time-lagged cross-correlations between COVID-19 new cases and each topic were calculated, using a 7-day moving average of both IOT values and COVID-19 confirmed cases and deaths to blunt the day-by-day fluctuation.

### Model Development and Assessment

Principal Components Analysis (PCA) was used to perform data dimensionality reduction, decrease the number of input variable, and filter out noisy or redundant information. For each country, two PCA models were calculated: one using unprocessed data and the other using a 7-day moving average smoothing. For the sake of comparability of the variables, PCA was applied to standardized data (i.e., with zero mean and unit variance). The PCA model was graphically inspected through PCA score and loading plots. PCA was assessed via 5-fold cross-validation, and the results obtained in each test sample were averaged. The amount of variance explained by each principal component (PC) in the model was inspected with scree plots, and, based on the elbow and Kaiser rules, the first two PCs (PC1 and PC2) were subsequently used for time-series modelling.^13^

Three different time series models were fitted on PC1 and PC2 values. The models tested were: Error Trend Seasonality (ETS), Autoregressive integrated moving average (ARIMA), and a feed-forward neural network autoregression (NNAR) model with one hidden layer.^14^ Models were fitted on a 30-day window and used to predict future PC1 and PC2 values up to 14 days. The fourteenth predicted day was aligned to the peak and base of each wave. The new data scores predicted with the time series models were then reinserted into the model as input variables.

For each country, the three models were compared in terms of ability to predict the PC1 and PC2 using the root-mean-square error (RMSE) of the predicted values. For each time-series model, the predictive ability of the models generated with raw data and 7-day moving average were compared. To further assess the PCA models based on both Google Trends data and conventional COVID-19 metrics, we also generated predictive models based on conventional COVID-19 metrics only; we then compared the predictive ability of models with and without Google Trends data by means of RMSE for each country.

### Web-application

An open-source web-application (https://predictpandemic.org) was developed in the R Shiny.^15^ Data are collected, imported, and updated daily for 188 countries from the sources mentioned above.

For web-application feature, specific countries, timeframe, and moving averages can be selected by the user. The web-application allows users to generate lineagraphs and streamgraphs to visualize IOT values and COVID-19 metrics, and to view worldwide trends over time in the form of a choropleth map. Relationships between the variables at the various lags can be explored with cross-correlations. The web-application allows fitting and evaluating PCA models, fitting a time-series model (either ETS or ARIMA), predicting PC components or any of the input variable of the model (including numbers of new cases and deaths), and evaluating the model performance graphically and with various metrics, such as RMSE and mean absolute error. The user has operational control on several model features, including the subset of variables to build the PCA model, the time window to fit the time-series model, and the time interval to predict.

## RESULTS

Three European countries (Italy, UK, and France), one Asian country (India), one Oceanian country (Australia), one North American country (US), one South American country (Brazil), and one African country (South African) were chosen as case studies. Cumulative number of cases and deaths in the selected countries are illustrated in Figure 1.

**Figure 1.**
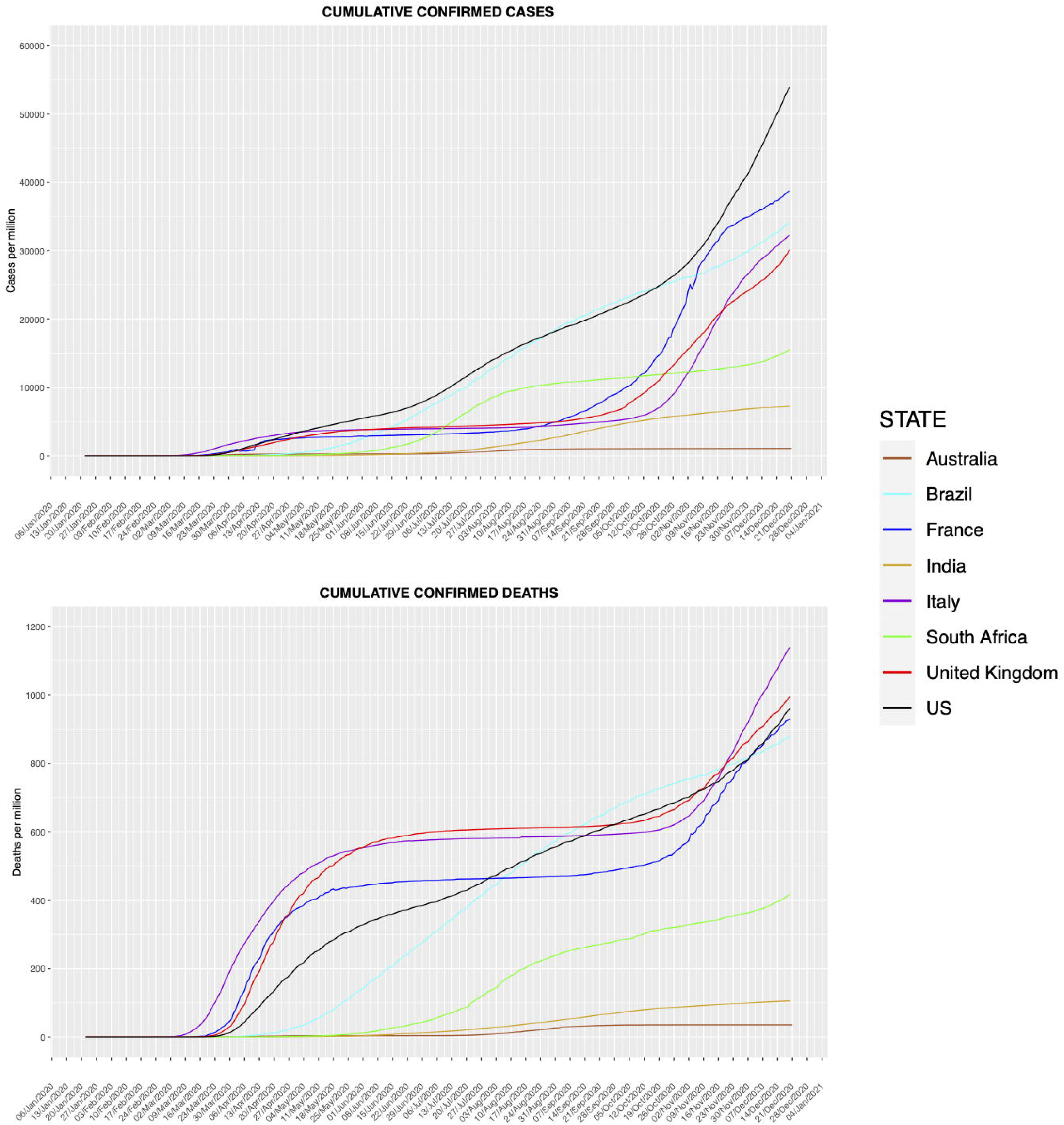
Cumulative number of confirmed cases (top panel) and deaths (bottom panel) per million for each country over time.

Table 1 summarizes information of the five-year analysis. Among the 20 screened topics, 13 showed seasonality, while the remaining were non-seasonal. Eleven searched topics showed a clear deviation from their five-year trend: ageusia, anosmia, chills, cough, eye pain, fever, headache, nasal congestion, rhinorrhea, shortness of breath, sore throat.

**Table 1.** Symptoms screened at the 5-year analysis

| **Table 1.** Symptoms screened at the 5-year analysis |  |  |
| --- | --- | --- |
| **Topic** | **Seasonality** | **Deviation from 5-year trend** |
| Abdominal Pain | Nonseasonal | No |
| Ageusia | Nonseasonal | Yes |
| Anorexia | Seasonal | No |
| Anosmia | Nonseasonal | Yes |
| Bone Pain | Nonseasonal | No |
| Chills | Seasonal | Yes |
| Conjunctivitis | Seasonal | No |
| Cough | Seasonal | Yes |
| Diarrhea | Seasonal | No |
| Eye Pain | Nonseasonal | Yes |
| Fatigue | Seasonal | No |
| Fever | Seasonal | Yes |
| Headache | Seasonal | Yes |
| Myalgia | Seasonal | No |
| Nasal Congestion | Seasonal | Yes |
| Nausea | Nonseasonal | No |
| Rhinorrhea | Seasonal | Yes |
| Shortness of Breath | Seasonal | Yes |
| Sore Throat | Seasonal | Yes |
| Tearing | Nonseasonal | No |
| Topic categorization into deviating and not-deviating from their 5-year trend was determined on the visual inspection of decomposition plots. |  |  |

The relationships between the number of new cases and each searched topic are illustrated in Supplementary Figures 1-8 Several symptoms, including ageusia, anosmia, cough, rhinorrhea, and sore throat were aligned with the COVID-19 epidemic in most countries and were searched on Google well before the number of COVID-19 confirmed cases peaked. On the other hand, other topics showed less evident variations (chills, eye pain) or deviated from their trend only during the first wave (headache, shortness of breath). Also, the peak of interest of all symptoms (except eye pain) anticipated that of confirmed COVID-19 cases in most countries, and topics increasing earlier reached their highest IOT value before those growing later. Similar patterns were observed IOT of searched terms were plotted over number of new confirmed deaths (Supplementary Figures 9-16).

The change of interest over time for all the topics is illustrated in Figure 2. Overall, the IOT values of the selected topics had a peak in March in all the selected countries. In Italy, France, South Africa, and, to a lesser extent, the UK and the US, there was a decrease in the searched terms after the first peak followed by a second peak. In India and Brazil, searches of medical terms remained high after the first peak, and no second peak was seen. In Australia, the IOT values of the selected topics returned to the pre-peak values soon after the first peak in March and remained low and stable.

**Figure 2.**
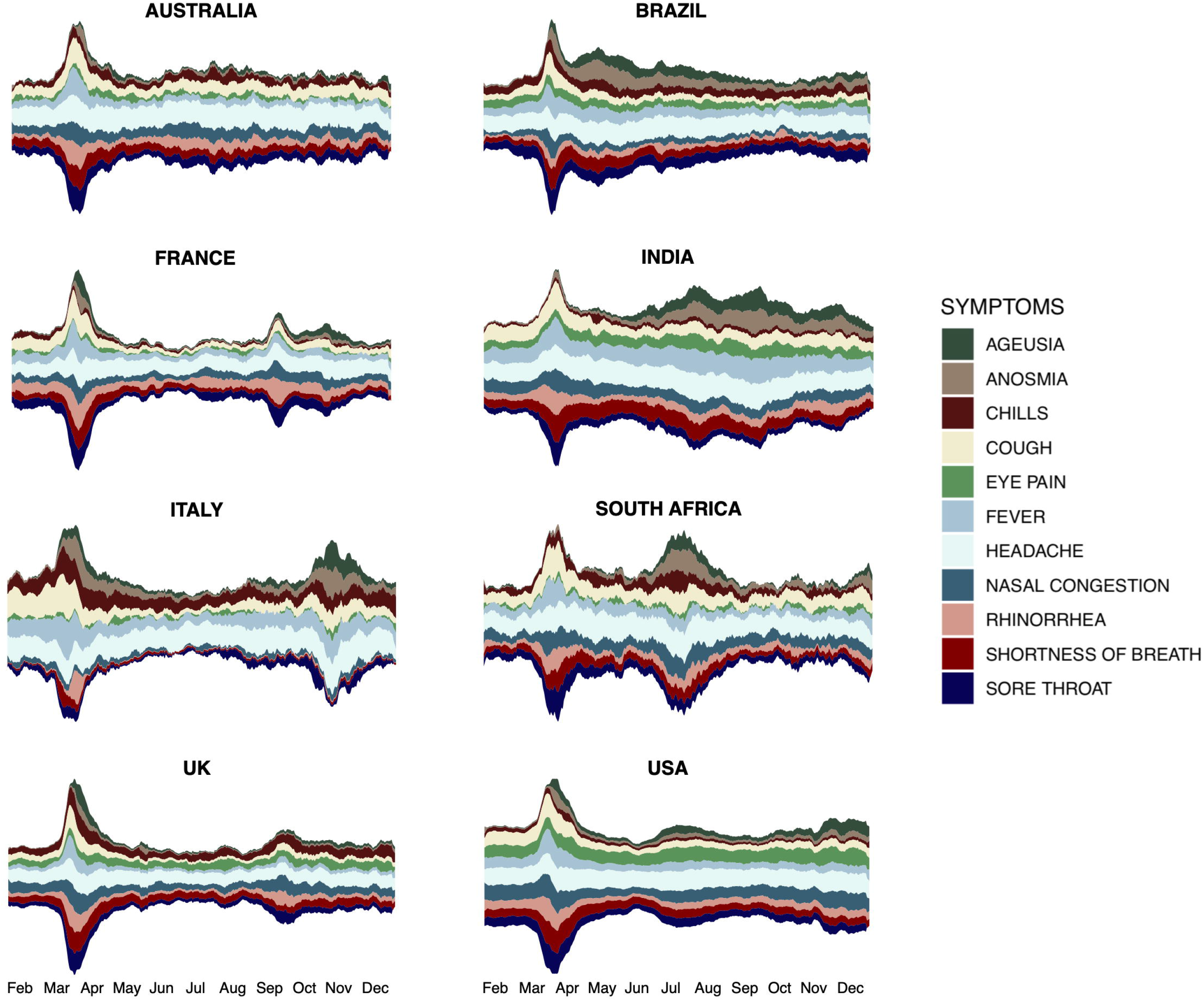
Streamgraphs of interest over time (IOT) index for each individual country. X-axis is given in months. IOT values were plotted as seven-day moving average.

Cross-correlations between each topic and the number of confirmed COVID-19 cases are reported in the Supplementary Tables 1-8. Overall, ageusia, anosmia, and headache were most consistently correlated with COVID-19 cases across the selected countries. The degree of correlation and the best time-lag largely varied as a function of the selected country and topic, but, overall, the optimal time-lag was 15 days.

The scores and loadings plots for the PCA models are given in Figures 3. The scores plot represents a summary of the collected data trends over time, while the loadings plot shows how strong each variable influences a PC. In the month of March 2020, all the selected countries deviated considerably from their previous scores and moved toward the PCs directions of the loadings of the Google searched terms, anticipating the increment in the number of searches of the symptoms related to COVID-19. The latest 14 days show a similar pattern for all the selected countries, with the latest days pointing toward the loadings’ directions of deaths and new cases. Specifically, for Italy, France, and South-Africa, the latest scores are in the same area of loading plots corresponding to deaths and new cases, indicating a stable trend in these metrics. On the other hand, the UK, the US, Brazil, and India followed a worsening pattern, as their scores kept moving toward the direction of new deaths and cases. Australia was the only of the selected countries showing an improving trend, with the scores points moving away from the loadings of COVID-19 deaths and new cases.

**Figure 3.**
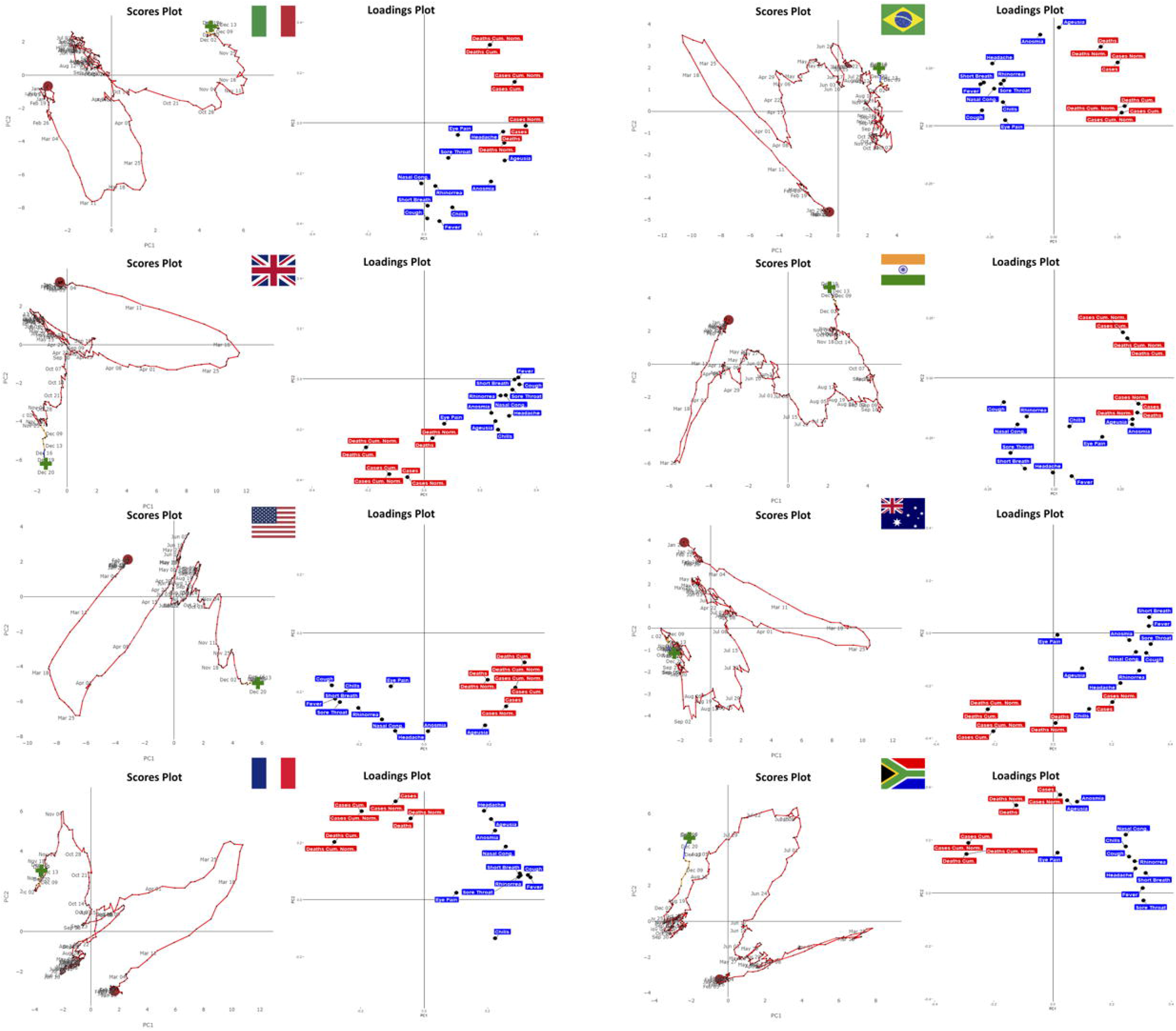
Principal component analysis scores and loadings plot for the selected countries.

Figure 4 illustrates the RMSE for the prediction of PC1 values with the 3 time-series models, in the various countries, using raw data and a 7-day moving average. Models based on the 7-day moving average lead to considerably smaller RMSE values as opposed to those calculated with raw data (median [IQR]: 0.74 [0.47-1.22] vs. 2.15 [1.55-3.89], respectively). Overall, predictions based on both searched terms and COVID-19 traditional metrics performed better than those not including Google searches (median [IQR]: 1.43 [0.74-2.36] vs. 1.78 [0.95-2.88], respectively), but the improvement in prediction varied as a function of the selected country and timeframe. Although ETS (median [IQR]: 1.44 [0.78-2.63]) led to slightly smaller RMSE than ARIMA (median [IQR]: 1.62 [0.93-2.50]) and NNAR (median [IQR]: 1.69 [0.97-3.03]) models, none of the tested time-series models clearly outperformed the remaining two, and the best model varied as a function of country, time range, and period of time selected. Similar results were obtained when trying to predict PC2 (Supplementary Figure 17).

**Figure 4.**
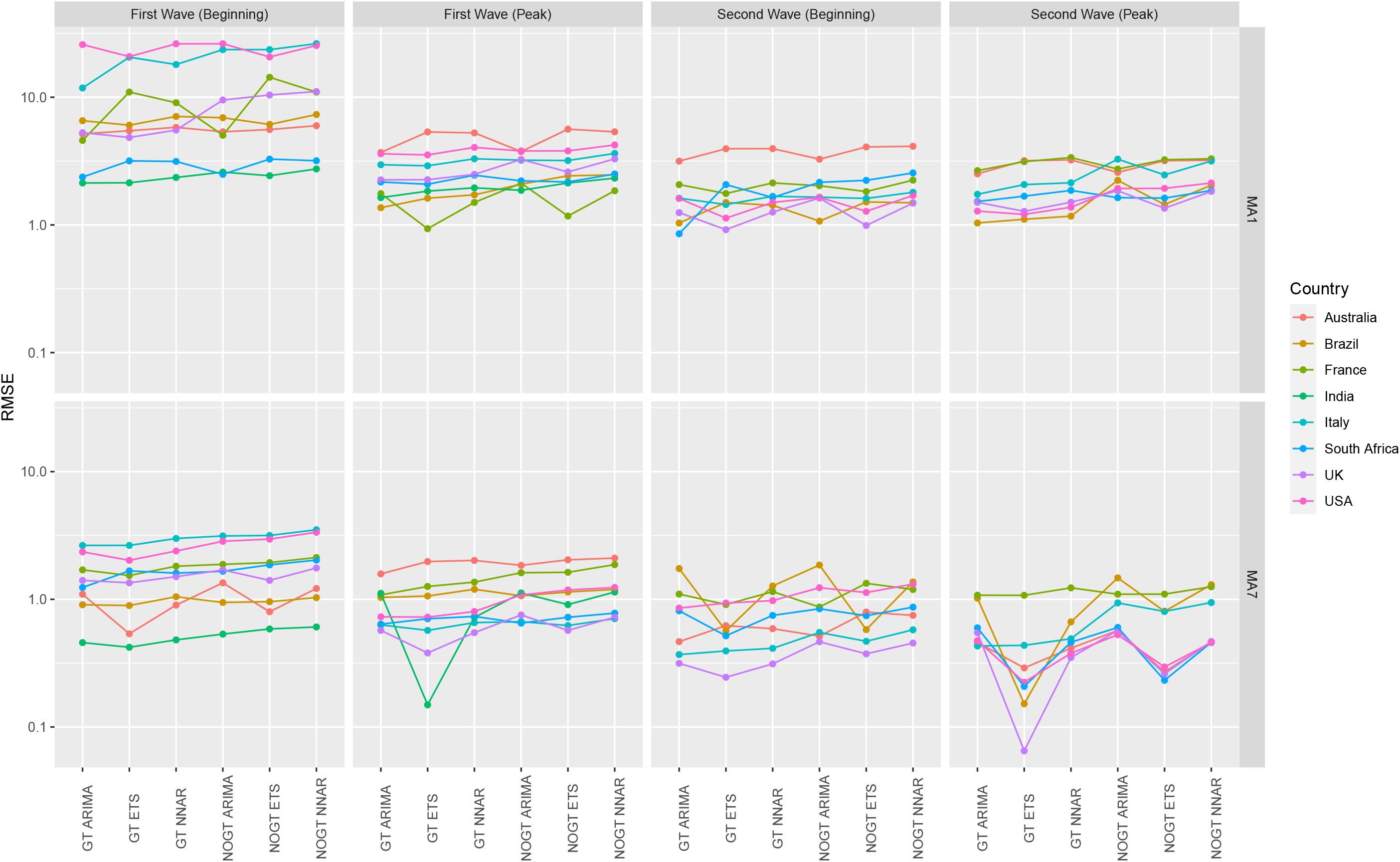
Root-mean-square errors (RMSE) of the prediction error for the principal component 2 of the various models for the selected countries. MA1 and MA7 indicate analyses performed on moving average of data 1 day (i.e., original data) and 7 days, respectively. GT indicates models based on both traditional COVID-19 metrics and Google Trends data, while NOGT models based on COVID-19 metrics only. ARIMA: Autoregressive integrated moving average; ETS: Error Trend Seasonality; NNAR: feed-forward neural network autoregression.

## DISCUSSION

In this study, we investigated the relationship between Google Trends searches of symptoms associated with COVID-19 and confirmed COVID-19 cases and deaths using PCA. We found that some of the searched terms showed an unusually high recent online interest that deviated considerably from their expected behavior and anticipated the peak of confirmed COVID-19 cases by days to weeks. This pattern was consistent across different countries and of similar magnitude. We developed and validated predictive models to forecast COVID-19 epidemic based on the combination of Google Trends searches of symptoms associated with COVID-19 and traditional COVID-19 metrics. We found that models incorporating Google Trends data performed generally better than those based solely on traditional COVID-19 metrics. We also developed a web-application (https://predictpandemic.org) to translate our approach into action.

Modeling epidemics is a complex task that depends on several assumptions and entails numerous limitations. The choice of data input is a crucial part of model development for accurate predictions.^16^ Most of the current models for COVID-19 rely on confirmed cases or deaths, but neither of these measures is satisfactory. Confirmed COVID-19 cases represent only a part of all infected subjects, as those with mild symptoms may not seek medical attention or get tested. Also, the number of confirmed cases is highly dependent on the number of tests performed, which varies greatly in different countries and in the diverse phases of the epidemic.

Confirmed COVID-19 deaths are likely to be a more reliable measure, but they occur in the final stages of the disease and are, therefore, a poor indicator to detect outbreaks at their earliest stages. Also, COVID-19 deaths are not uniformly reported among the different countries and may vary as a function of the healthcare systems, population demographic and public health status.

In the past decade, there has been an increasing interest in the use of internet big data to understand patterns of disease, population behaviors, and make surveillance of infectious disease. Despite being initially welcomed with enthusiasm, models based only on Google Trends data proved to be not accurate in determining the absolute numbers of cases in epidemics, but were helpful in identifying temporal dynamics, anticipating peaks, and improving forecasting when used in conjunction with (and not in place of) traditional data.^17,18^

Our study identified patterns of Google searches of several symptoms and signs associated with COVID-19 in a consistent way across the studied countries. Overall, Google searches of COVID-19 symptoms followed a similar trend to that of the COVID-19 epidemic, and, importantly, anticipated traditional COVID-19 metrics. This behavior can contribute to early recognition of new waves and epidemic peaks.

The interpretation of symptom search behavior during COVID-19 outbreaks should be carefully considered. Dynamics of online searches may show atypical patterns during pandemics where major restrictions occur, including shutdowns of economic activities, movement restrictions, and healthcare overload.^19^ Constant media attention may contribute to raising the interest for some of the studied topics.^20^ COVID-19 received extensive coverage that might have precipitated unusually high interest during lockdowns.^21^ That is, our findings on online search behavior might be secondary to general media interest in specific COVID-19 symptoms, rather than a primary, and possibly predictive, consequence of COVID-19 sufferers researching their own symptoms, in real time. All the selected countries had a peak in searches of medical terms in or around March and April 2019, including those countries with low numbers of cases at that time, such as South Africa and India. This pattern may indicate that curiosity and media clamor toward the new pandemic can explain part of the first peak in searched terms, in agreement with a previous study by Szmuda and colleagues.^22^ After the first peak, however, Google search behavior followed different patterns across countries, and resembled the course of the COVID-19 epidemic. In those countries having a second wave, such as Italy, France, the UK, or the US, the number of Google searches had a second peak; however, the height of second peak of the searches was lower than that of first peak, despite a higher number of reported cases and deaths, suggesting that the first peak in searched terms could have been inflated by the individual curiosity toward the new pandemic. South Africa had a second peak in its searches in July 2019, when the country had its first wave. In India, the first peak in searches was followed by a steady increase from June 2019, remained stable until October 2019, and then decreased gradually, resembling the shape of COVID-19 epidemic in this country. In Australia, who effectively managed the COVID-19 epidemic and had among the lowest infection and death rates in the world, the IOT of the various searches terms after the first peak remained low and comparable to pre-peak values.

We observe that not all the selected topics reached their peak searches at the same time, but they had different time patterns, which were fairly consistent across all countries. We believe that the intense and simultaneous media coverage of all the selected topics should have had the same effect at the same date if this search behavior were entirely caused by the media influence.^23^ Ageusia and anosmia showed the highest correlations when lagged by a few days, while cough, fever, nasal congestion, sore throat, rhinorrhea, and shortness of breath anticipated COVID-19 by up to two weeks. This finding is consistent with the clinical course of COVID-19; in a large multicenter European study, olfactory and gustatory dysfunctions were among the latest and first manifestations in approximately 65% and 12% of patients, respectively.^10^

Beside describing how google search terms changed over time in different countries and investigating their relationship with the numbers of cases and deaths, we also developed models combining interest-over-time values of searches of COVID-19 symptoms with traditional metrics (e.g., number of new cases, number of new deaths) to predict the course of COVID-19 epidemic, and we compared the prediction ability of these models against that of models based only on traditional metrics. The use of the PCA approach allowed us to reduction dimensionality, summarizing information into 2 PCs and filtering out the noisy or redundant information. Another advantage of PCA was to provide visual representations of data patterns, similarity trends, and outliers. The PCA approach is highly flexible, and potentially allows accommodating new variables of interest in future versions of our application. As the PCA itself does not make any predictions, we processed the PC computed values with different time-series models (i.e., ETS, ARIMA and NNAR), enabling us to extract the most useful information from the variability of the data to make predictions. As new data scores predicted with the time series models were reinserted into the PCA model, our approach allows extracting the predicted values of any of the input variable, including number of new cases and deaths. Models integrating IOT values of the searched topics and COVID-19 traditional metrics generally outperformed models based solely on confirmed cases and deaths, leading to improved predictions. There was no single best model in this study, and the best performing time series model varied as a function of the country, time frame, and moving average. Predictions were more accurate, leading to considerably smaller RMSE when obtained on a 7-day moving average, rather than on daily data. This result is not surprising as Google Trends data have high daily fluctuations, and COVID-19 reported cases greatly oscillate reflecting testing and reporting practices and contingencies.^24,25^

To translate our results into practice so that the scientific community, agencies, and even curious users could potentially use them, we developed a web-application, freely available at https://predictpandemic.org. The application is interactive and updates the data on a daily basis, so it operates in nearly real-time. It allows the user to visualize data for 188 world countries choosing any time frame.

Also, COVID-19 traditional metrics and google search terms IOT over time can be visualize globally on different graphs. The application the user to explore cross-correlations among selected keywords, to generate predictive models with default or a user-selected subset of variables, and to check model performance.

Another potential use of Google Trends is the opportunity to filter the results based on precise geographic locations having more granular data. These filters include nations, but also states, regions, and even cities in some areas. As not all areas of a country are equally affected nor have the same risk of having COVID-19 outbreaks, this function may allow regional predictions. We are currently planning to provide more granular data in our web-application.

The present study has limitations. The Google Trends algorithm is a ‘black box’, and the exact calculation formula for the interest over time and raw data have never been made public. Searched results may differ slightly when download by different computers or on different days, although we conducted search-research reliability, which showed excellent reliability for most of the topics included in this study (data not shown). The exclusion of those symptoms with no significant deviation from their five-year trends reduced the possibility of spurious correlations, but it was not possible to account for seasonality in the selected topics. In other words, a small proportion of the increasing trend in some topics might be explained by their usual seasonal variations. The results of this study may not apply to countries where Google is not a popular search engine or where Google is censored or limited in its use. However, this approach can be applied to other search engines (e.g., Baidu, Yahoo, Naver), as was done in previous studies on different diseases and on COVID-19 in Hubei province, China.^26,27^ Geographical areas and groups of people (elders and children) with scarce Internet access cannot be studied with this strategy, and our results may not apply to largely rural countries. This study included only the most common clinical manifestations of COVID-19, and only a few selected countries were included as a case study. However, information and models for every country can be found on our web-application.

Future work will include increased data granularity allowing to have information and make predictions at a regional level. Also, other metrics of interest, such as hospitalizations, will be included in our analysis as outcomes. Finally, we are planning to allow the user to generate a one-page report for each individual country, summarizing the most relevant information.

In conclusion, the results of this study show that Google Trends based on online searches during the COVID-19 pandemic may anticipate outbreaks by up to two weeks. The inclusion of digital online searches in statistical models may improve the nowcasting and forecasting of COVID-19 epidemic, and could be used as one of the surveillance systems employed by government agencies and supranational organizations to refine their monitoring of COVID-19 disease. We provide a free web-application operating with nearly real-time data that can be used by any user to make predictions of outbreaks, improve estimates of dynamics of ongoing epidemics, and anticipate future or rebound waves.

## Data Availability

Used data are freely available at the following website: https://predictpandemic.org

