## Supplementary Appendix for "Forecasting the COVID-19 epidemic integrating symptom search behavior: an infodemiology study"

### Supplementary information for term searches on Google Trends

Terms on Google Trends were searched using the «topic» function. In specific cases, Google Trends indicates the category of the topic, which is provided below between brackets for each term search included in the analysis:

- abdominal pain (syndrome)
- ageusia (topic)
- anorexia (symptom)
- anosmia (topic)
- bone pain (disease)
- chills (topic)
- conjunctivitis (topic)
- cough (topic)
- diarrhea (topic)
- eye pain (topic)
- fatigue (medical condition)
- fever (medical condition)
- headache (medical condition)
- myalgia (topic)
- nasal congestion (syndrome)
- nausea (disorder)
- rhinorrhea (medical condition)
- shortness of breath (disease)
- sore throat (topic)
- tearing (topic)

Supplementary Figure 1. Line graphs showing the interest over time (IOT) of the selected topic searches (blue lines) and their relationship with the normalized number of incident cases per million people (histogram) in Australia. Data are plotted as a 7-day moving average to smooth day-by-day fluctuations.

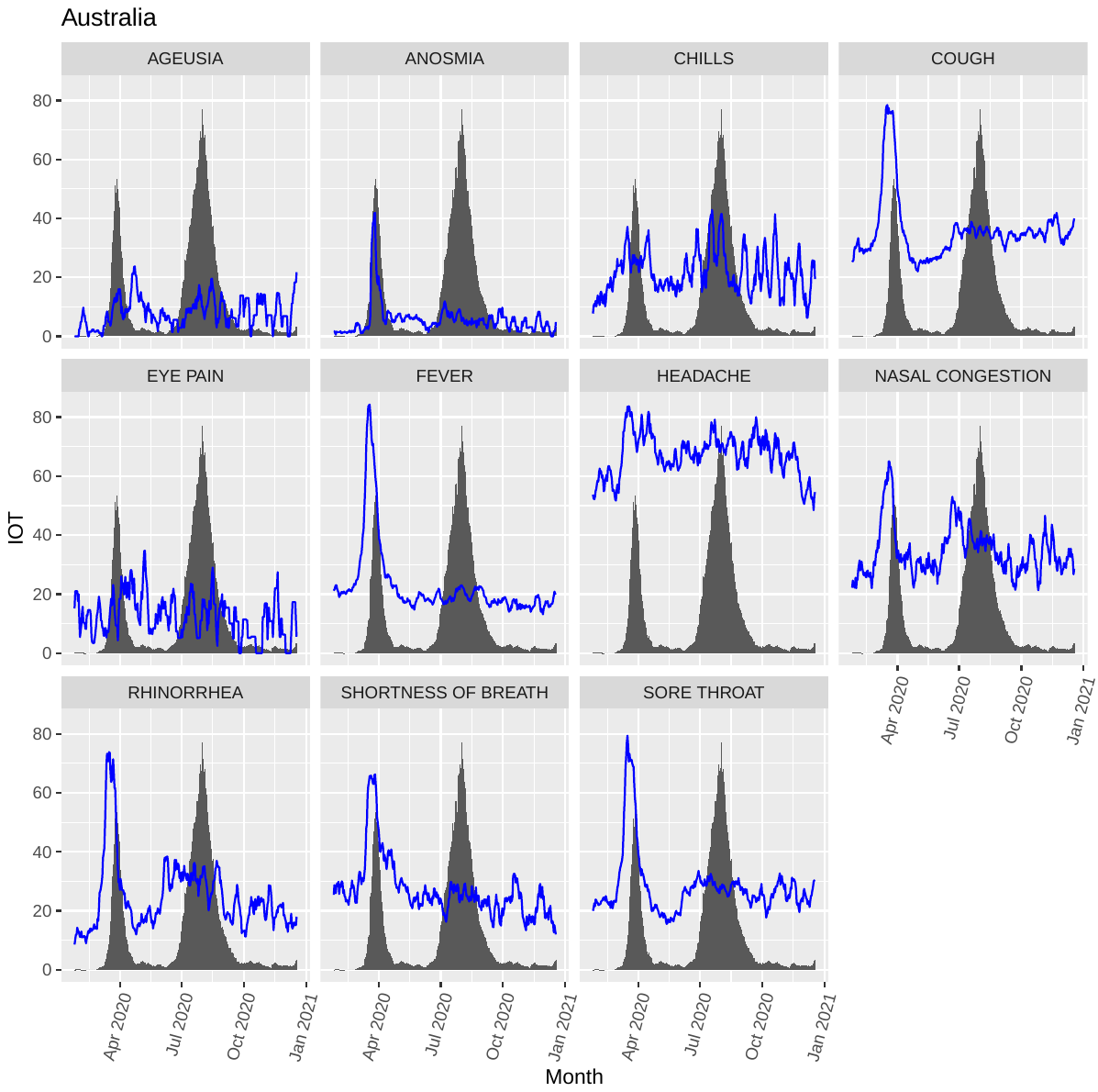

Supplementary Figure 2. Line graphs showing the interest over time (IOT) of the selected topic searches (blue lines) and their relationship with the normalized number of incident cases per million people (histogram) in Brazil. Data are plotted as a 7-day moving average to smooth day-by-day fluctuations.

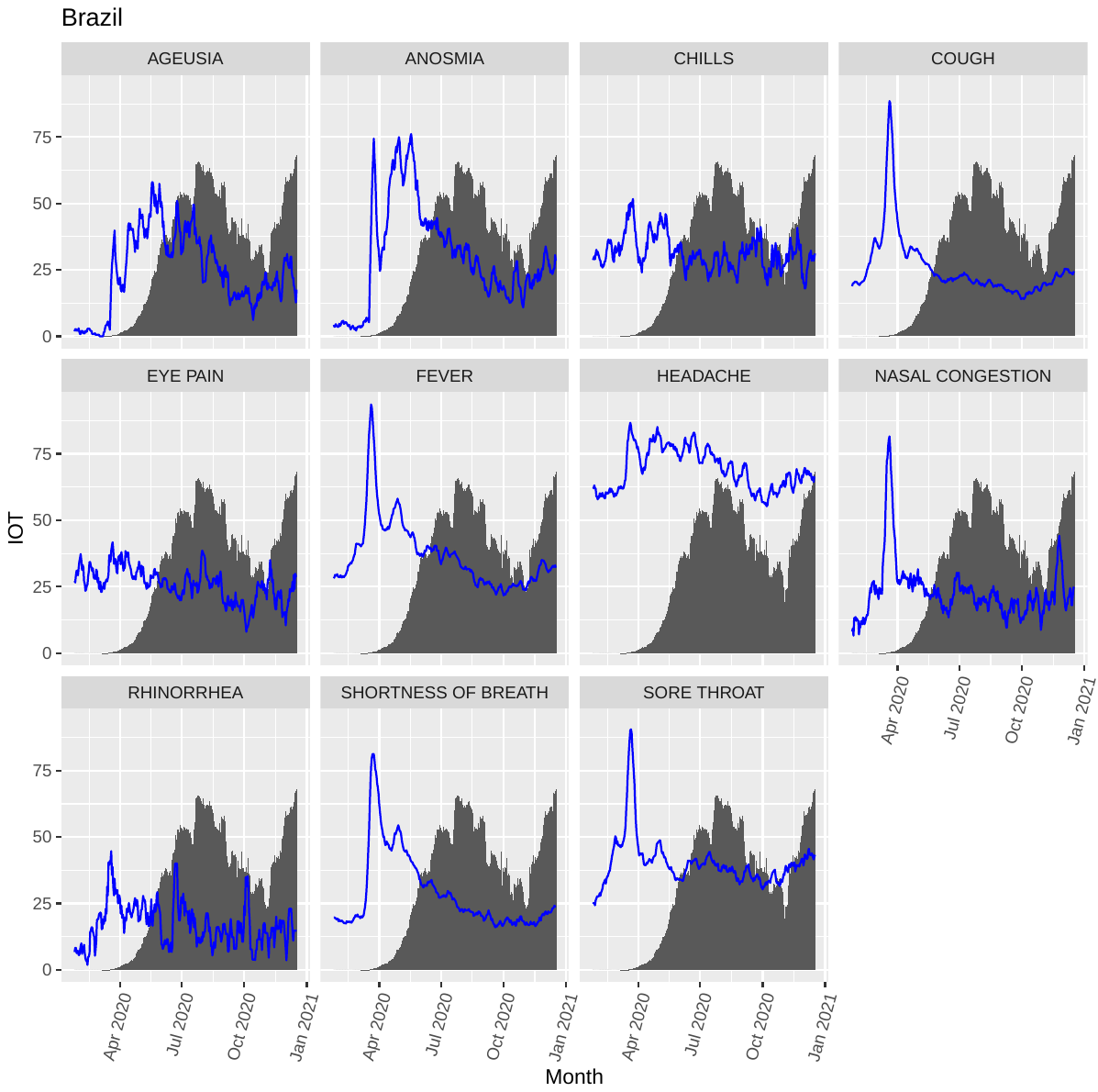

Supplementary Figure 3. Line graphs showing the interest over time (IOT) of the selected topic searches (blue lines) and their relationship with the normalized number of incident cases per million people (histogram) in France. Data are plotted as a 7-day moving average to smooth day-by-day fluctuations.

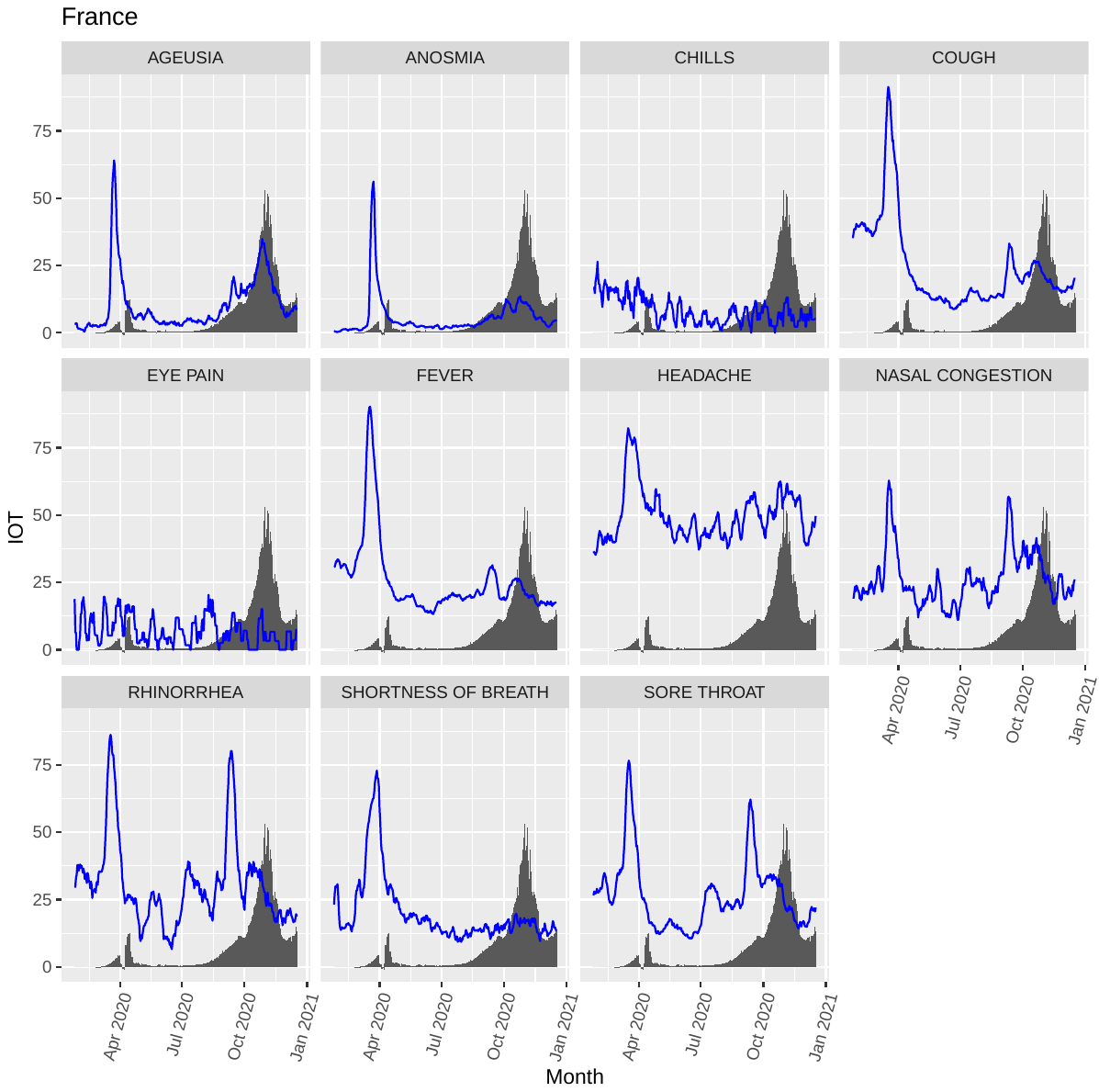

Supplementary Figure 4. Line graphs showing the interest over time (IOT) of the selected topic searches (blue lines) and their relationship with the normalized number of incident cases per million people (histogram) in India. Data are plotted as a 7-day moving average to smooth day-by-day fluctuations.

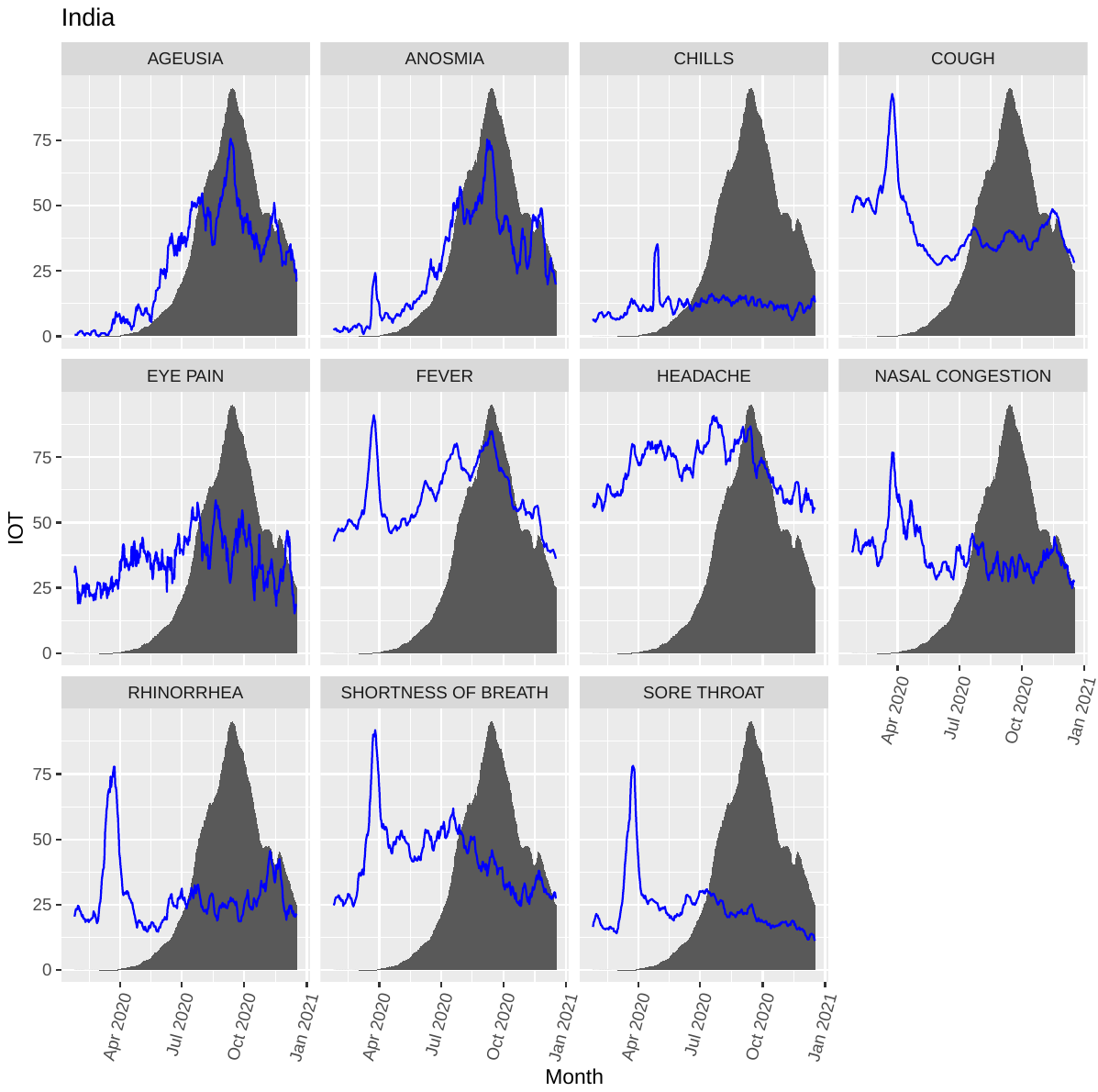

Supplementary Figure 5. Line graphs showing the interest over time (IOT) of the selected topic searches (blue lines) and their relationship with the normalized number of incident cases per million people (histogram) in Italy. Data are plotted as a 7-day moving average to smooth day-by-day fluctuations.

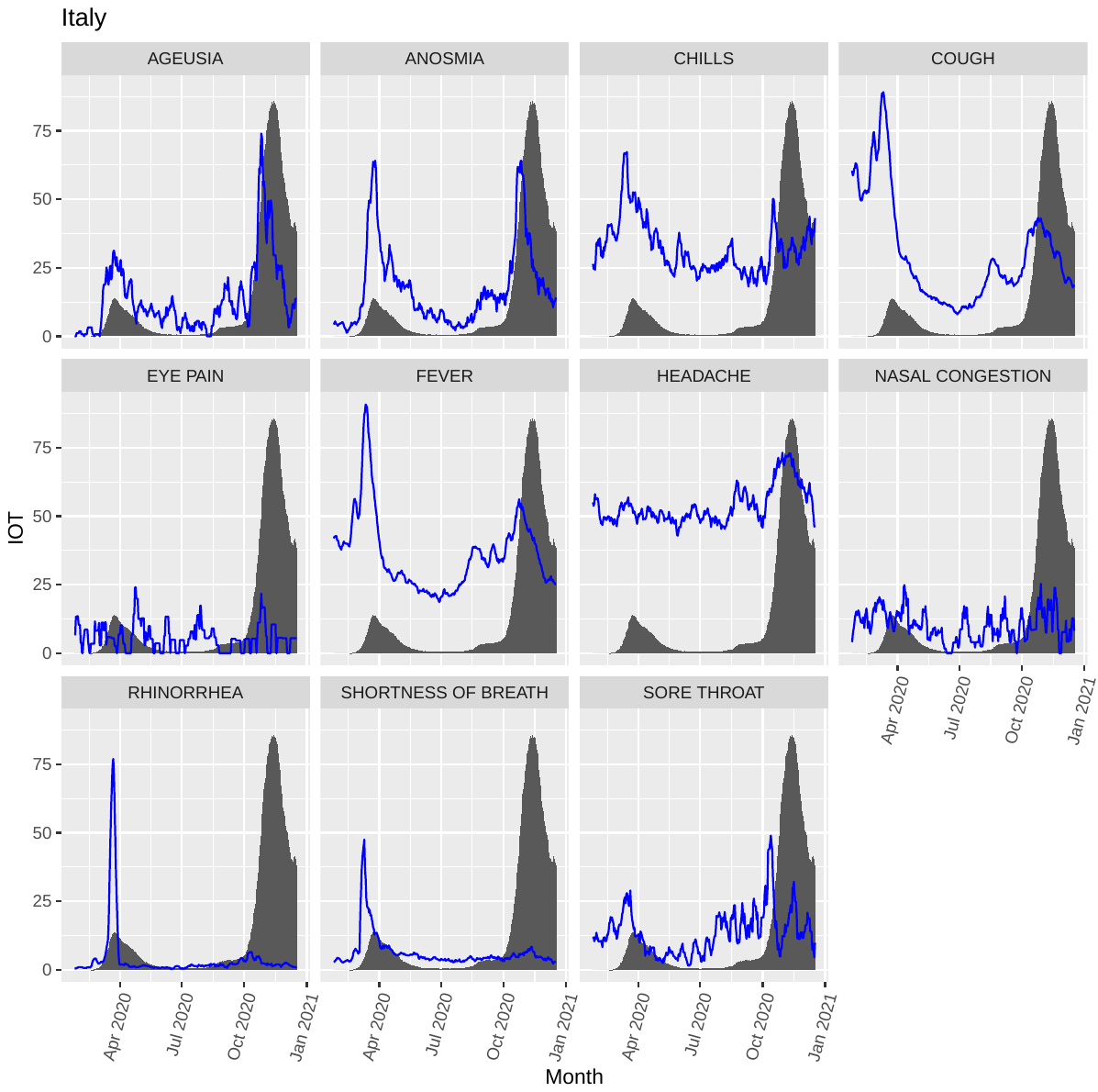

Supplementary Figure 6. Line graphs showing the interest over time (IOT) of the selected topic searches (blue lines) and their relationship with the normalized number of incident cases per million people (histogram) in South Africa. Data are plotted as a 7-day moving average to smooth day-by-day fluctuations.

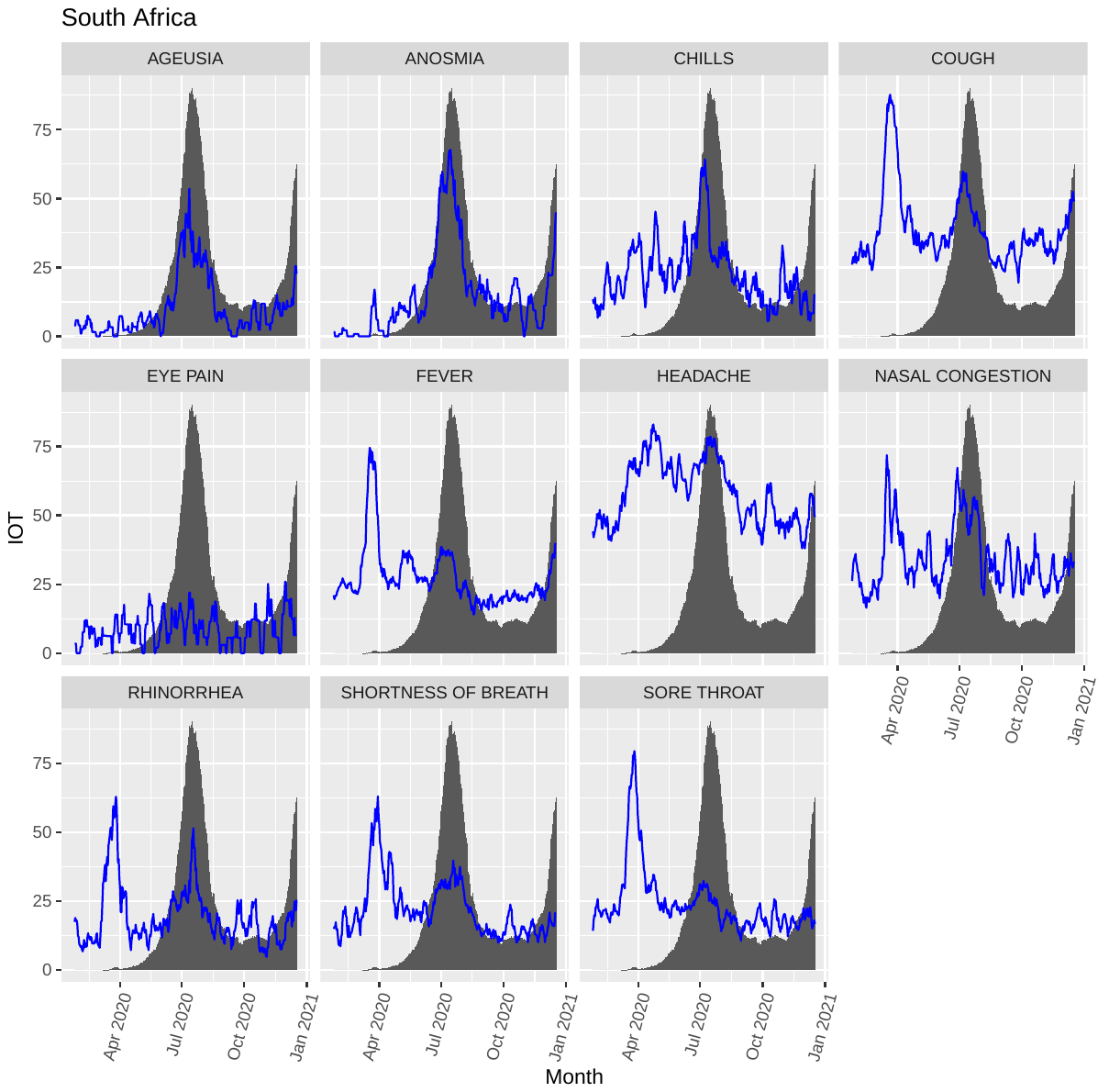

Supplementary Figure 7. Line graphs showing the interest over time (IOT) of the selected topic searches (blue lines) and their relationship with the normalized number of incident cases per million people (histogram) in the United Kingdom. Data are plotted as a 7-day moving average to smooth day-by-day fluctuations.

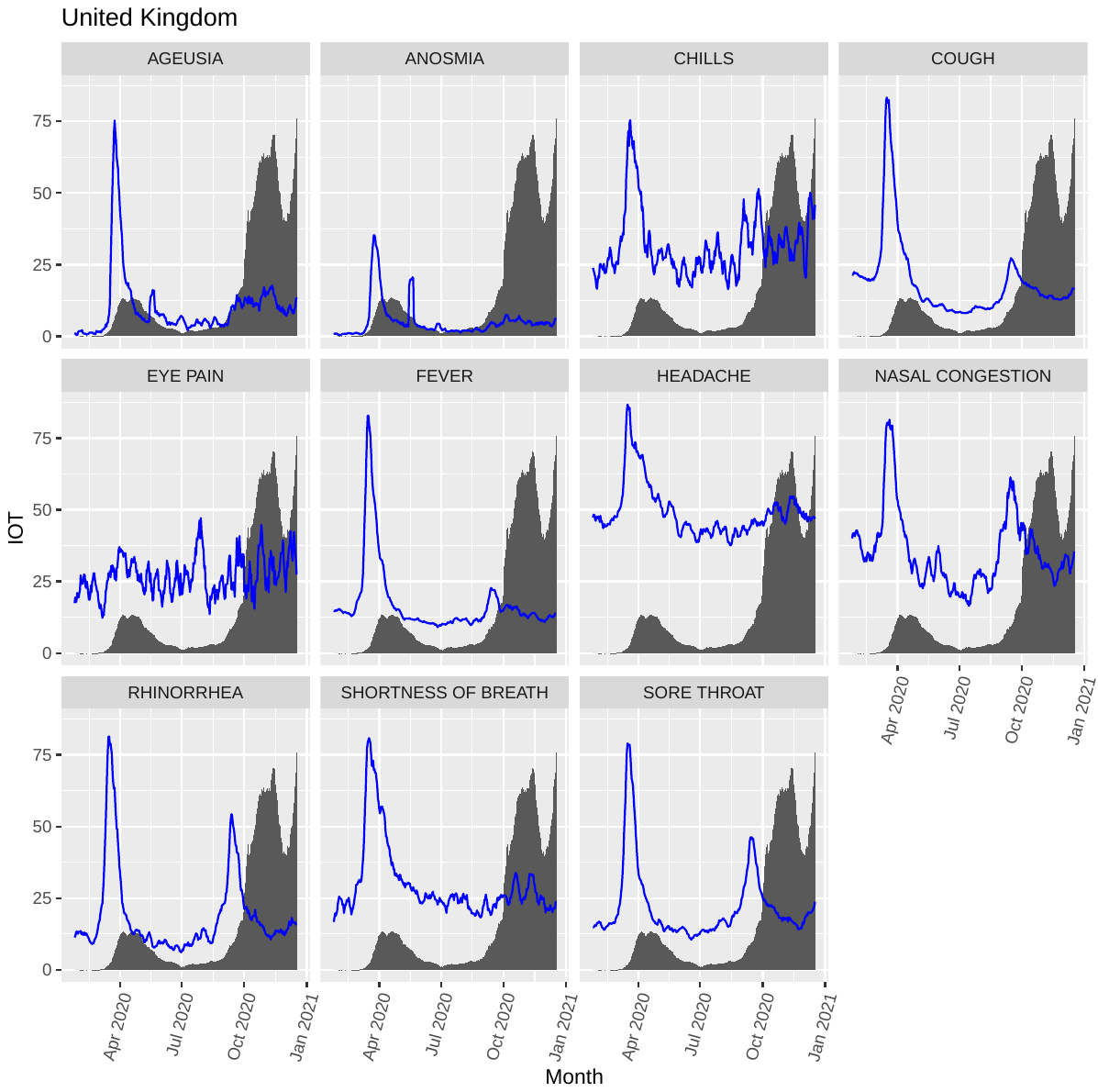

Supplementary Figure 8. Line graphs showing the interest over time (IOT) of the selected topic searches (blue lines) and their relationship with the normalized number of incident cases per million people (histogram) in the US. Data are plotted as a 7-day moving average to smooth day-by-day fluctuations.

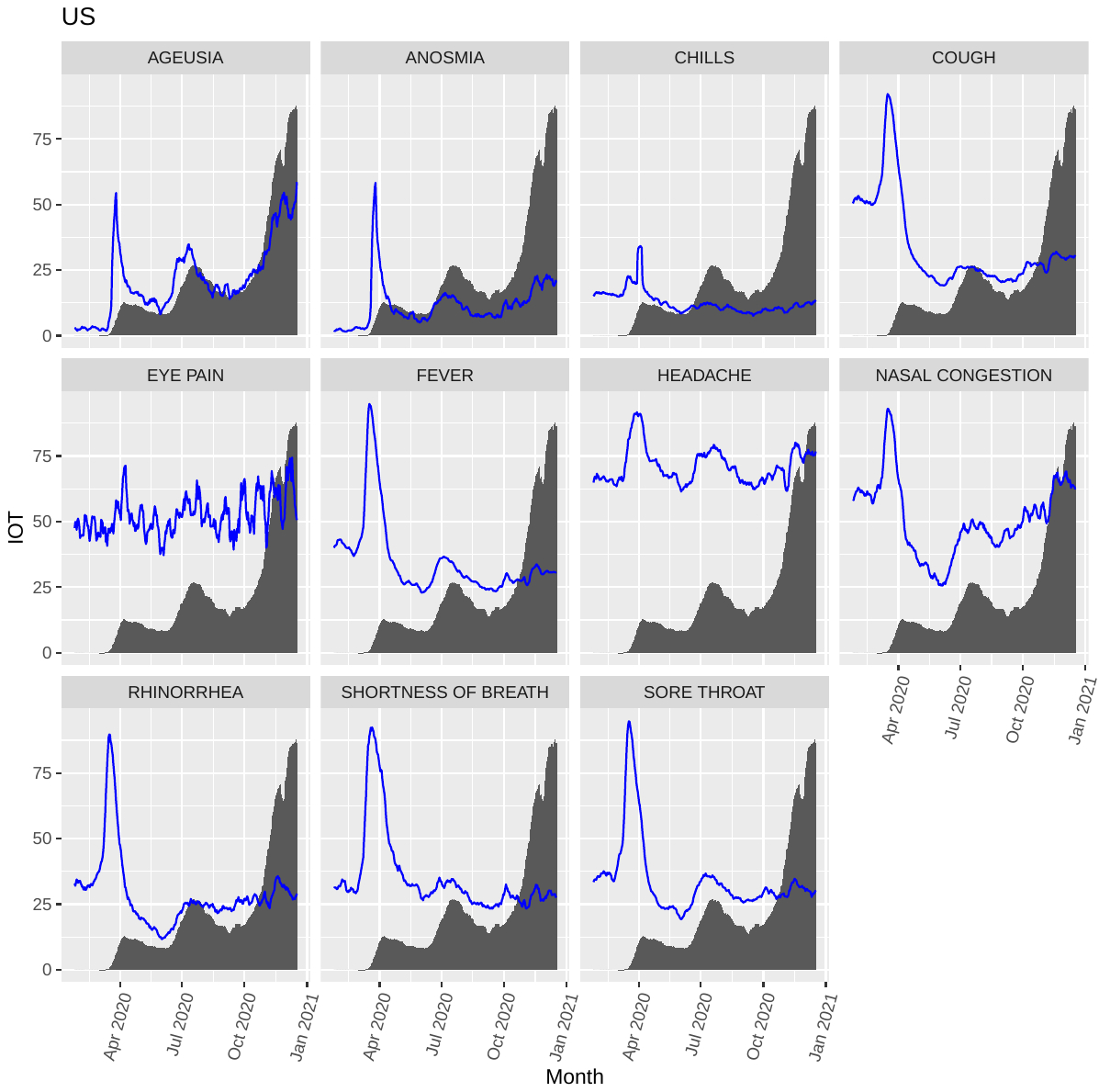

Supplementary Figure 9. Line graphs showing the interest over time (IOT) of the selected topic searches (blue lines) and their relationship with the normalized number of incident deaths per million people (histogram) in Australia. Data are plotted as a 7-day moving average to smooth day-by-day fluctuations.

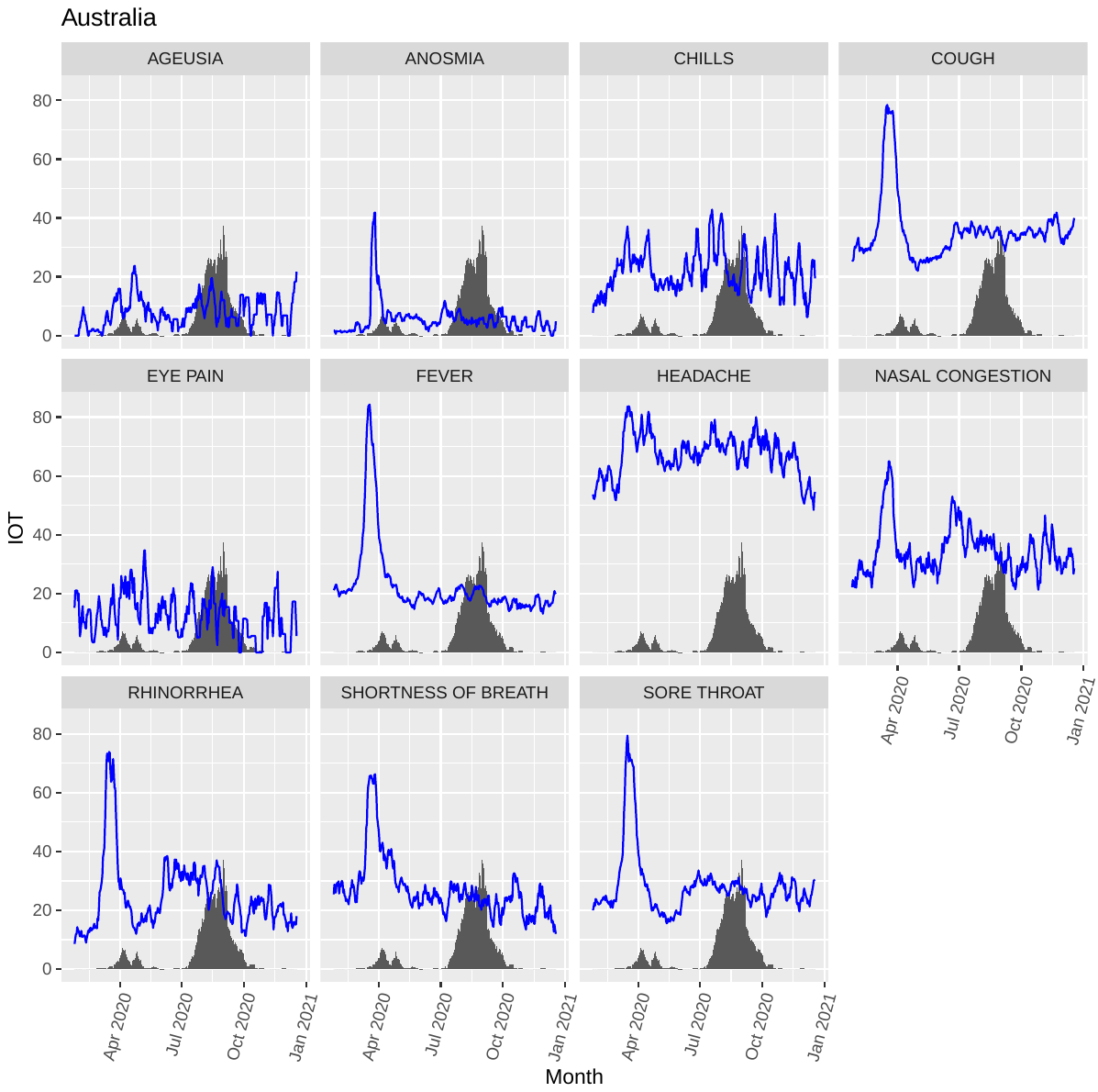

Supplementary Figure 10. Line graphs showing the interest over time (IOT) of the selected topic searches (blue lines) and their relationship with the normalized number of incident deaths per million people (histogram) in Brazil. Data are plotted as a 7-day moving average to smooth day-by-day fluctuations.

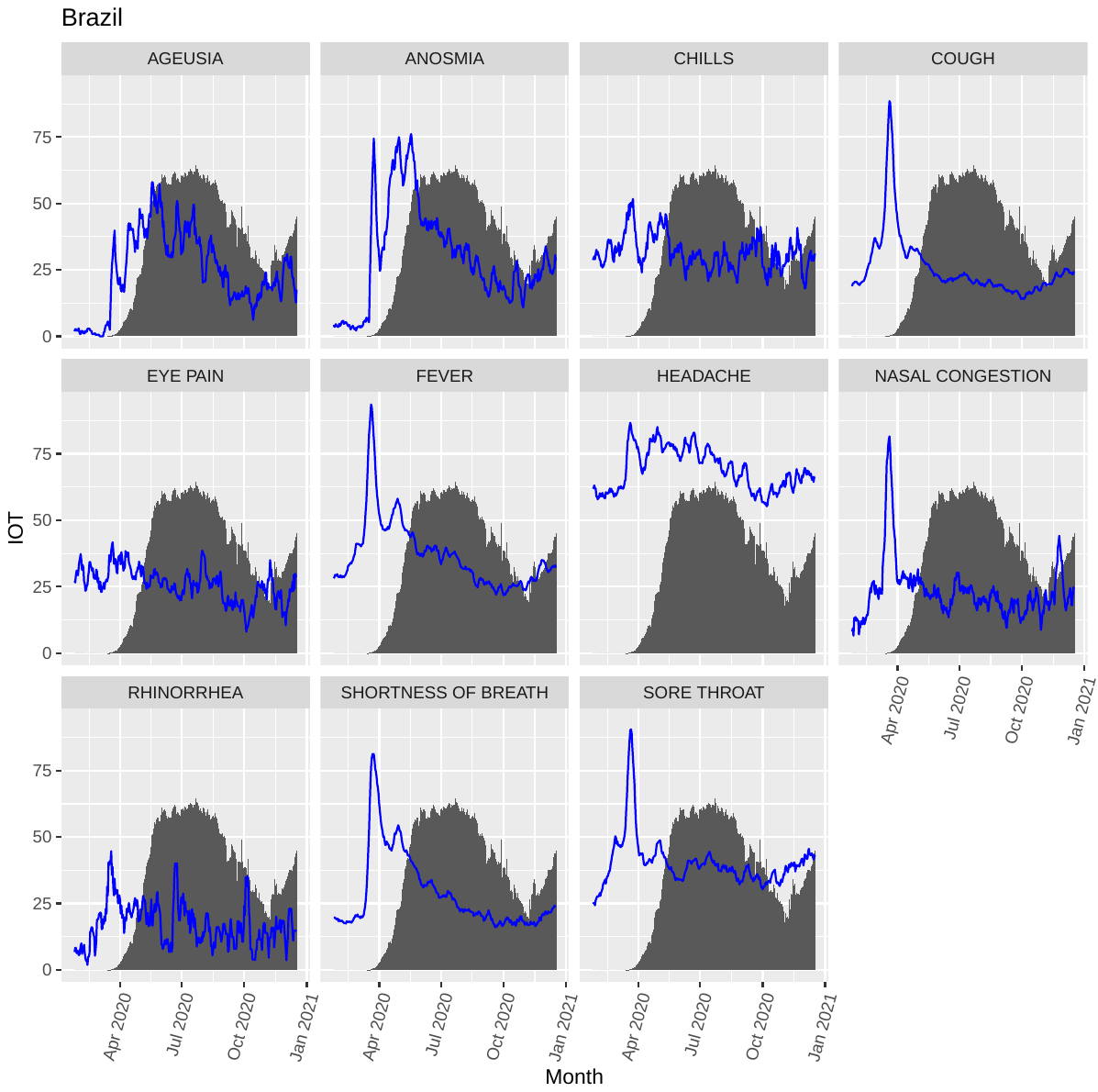

Supplementary Figure 11. Line graphs showing the interest over time (IOT) of the selected topic searches (blue lines) and their relationship with the normalized number of incident deaths per million people (histogram) in France. Data are plotted as a 7-day moving average to smooth day-by-day fluctuations.

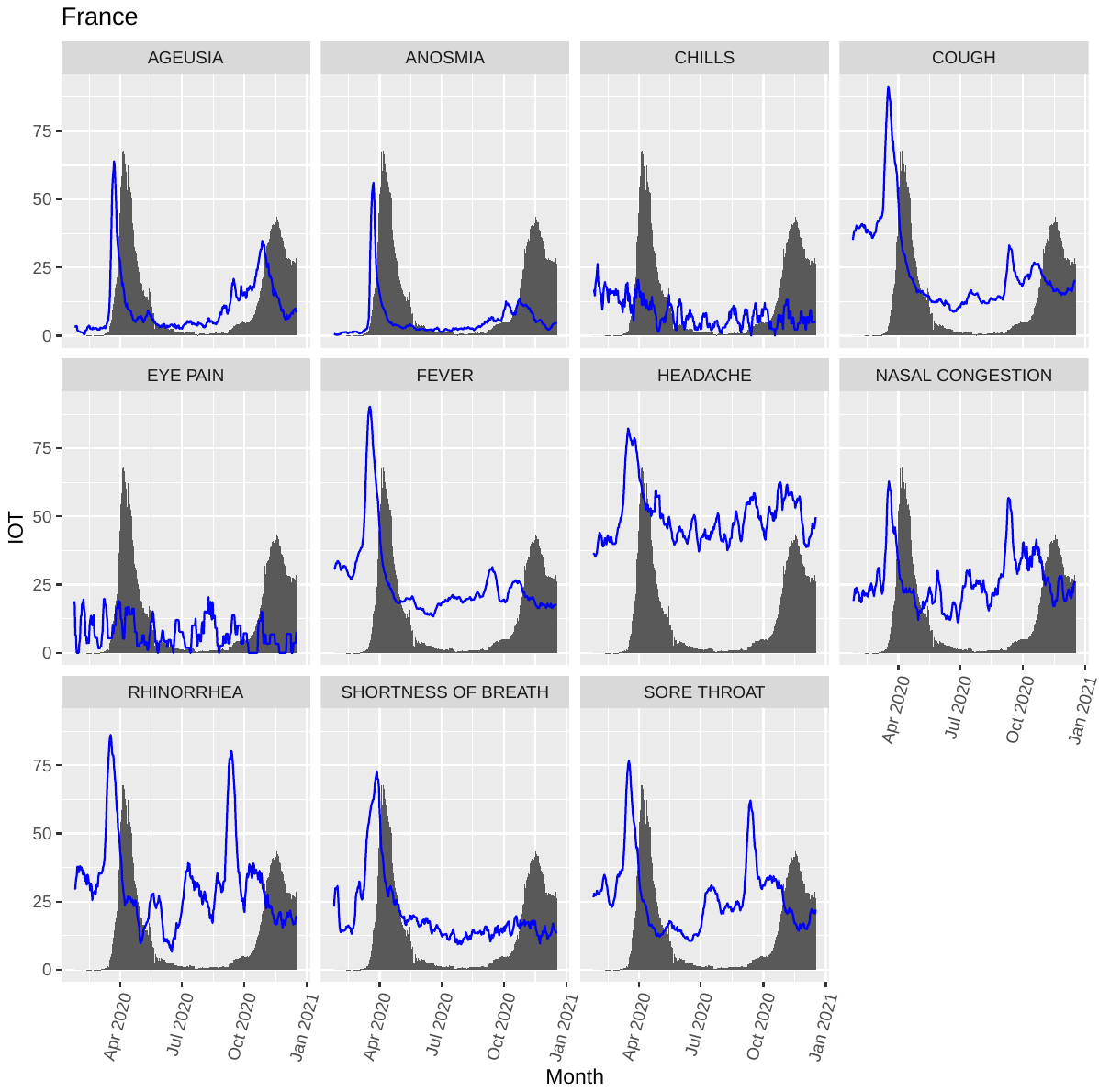

Supplementary Figure 12. Line graphs showing the interest over time (IOT) of the selected topic searches (blue lines) and their relationship with the normalized number of incident deaths per million people (histogram) in India. Data are plotted as a 7-day moving average to smooth day-by-day fluctuations.

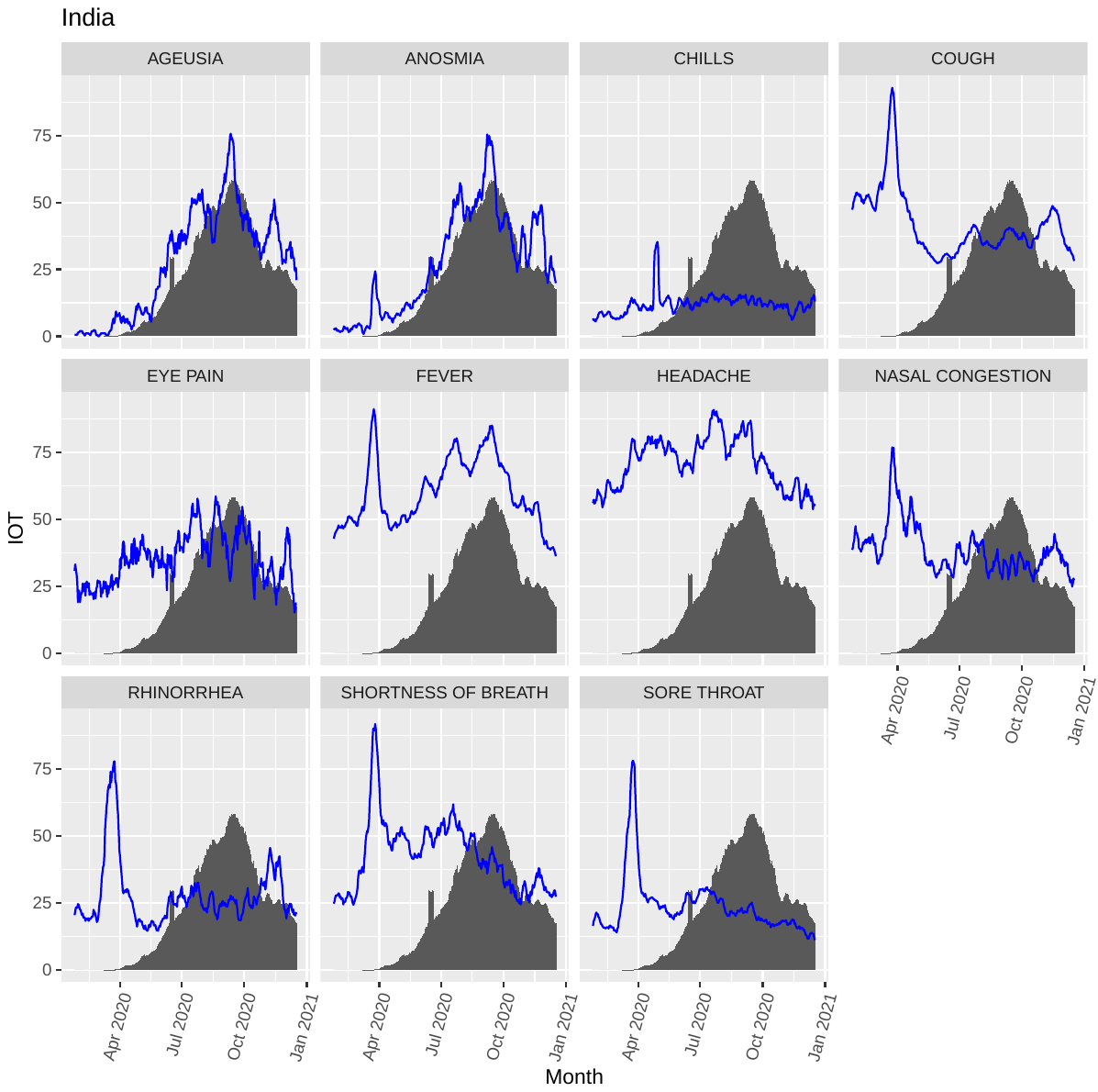

Supplementary Figure 13. Line graphs showing the interest over time (IOT) of the selected topic searches (blue lines) and their relationship with the normalized number of incident deaths per million people (histogram) in Italy. Data are plotted as a 7-day moving average to smooth day-by-day fluctuations.

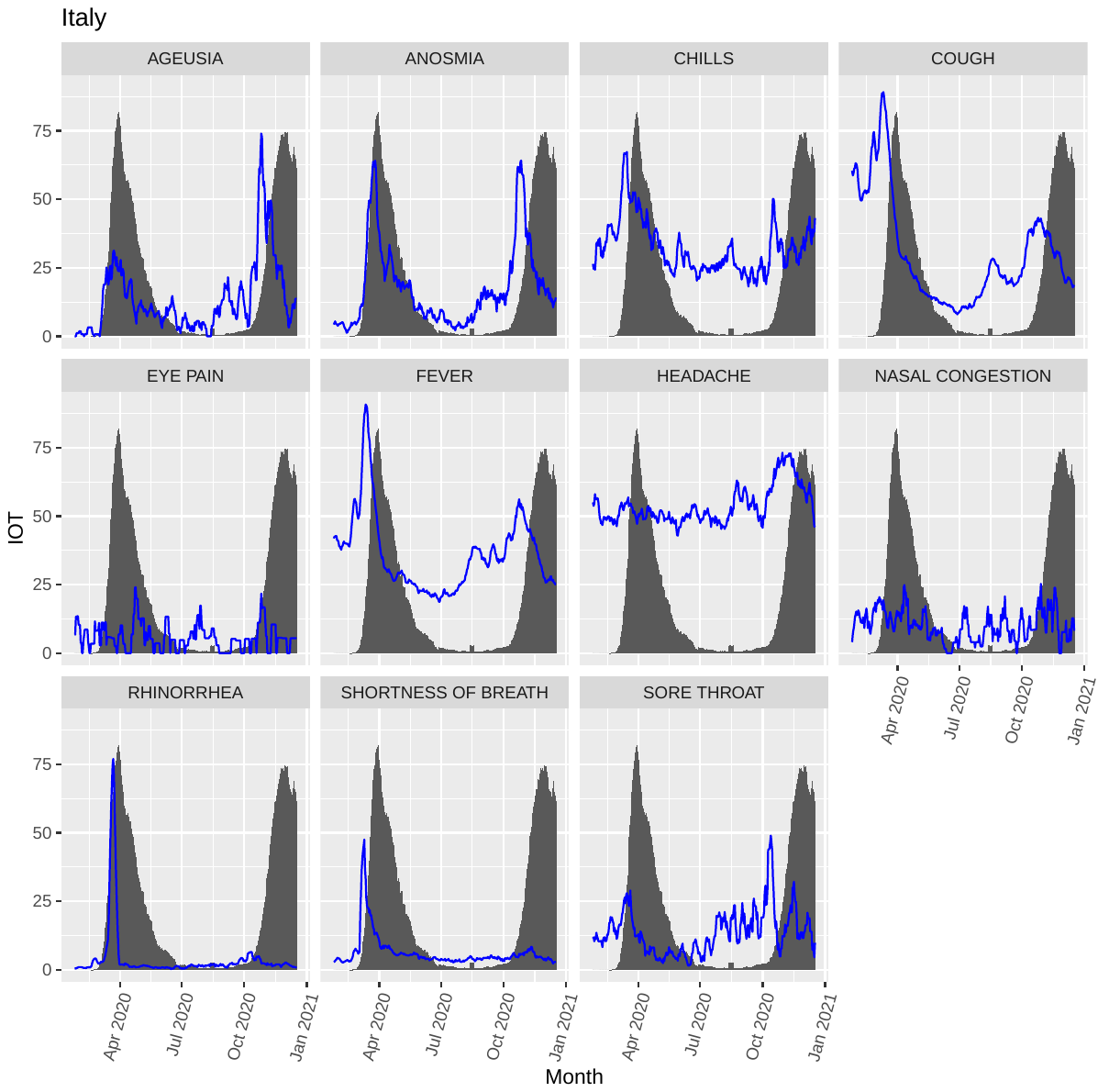

Supplementary Figure 14. Line graphs showing the interest over time (IOT) of the selected topic searches (blue lines) and their relationship with the normalized number of incident deaths per million people (histogram) in South Africa. Data are plotted as a 7-day moving average to smooth day-by-day fluctuations.

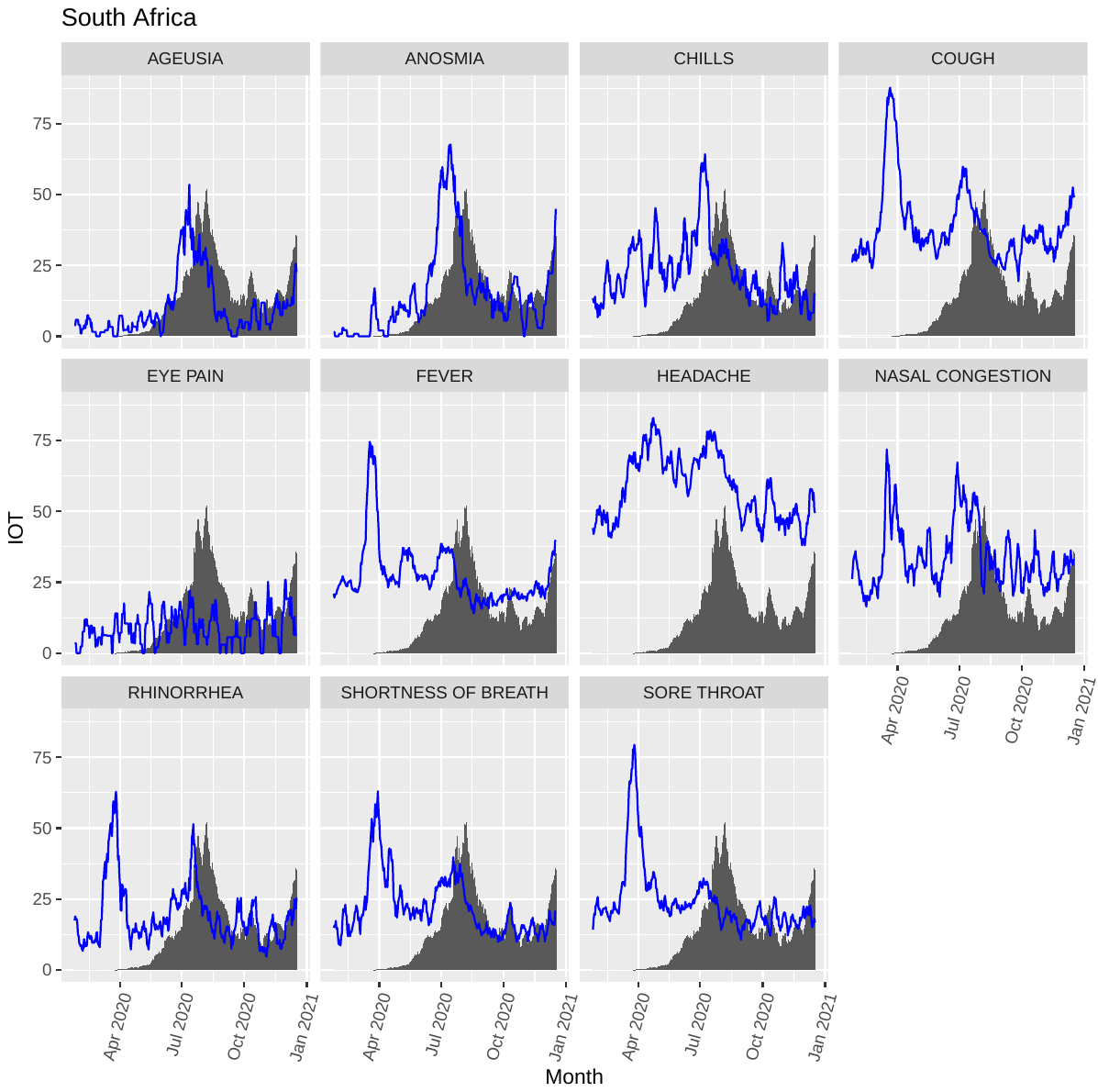

Supplementary Figure 15. Line graphs showing the interest over time (IOT) of the selected topic searches (blue lines) and their relationship with the normalized number of incident deaths per million people (histogram) in the United Kingdom. Data are plotted as a 7-day moving average to smooth day-by-day fluctuations.

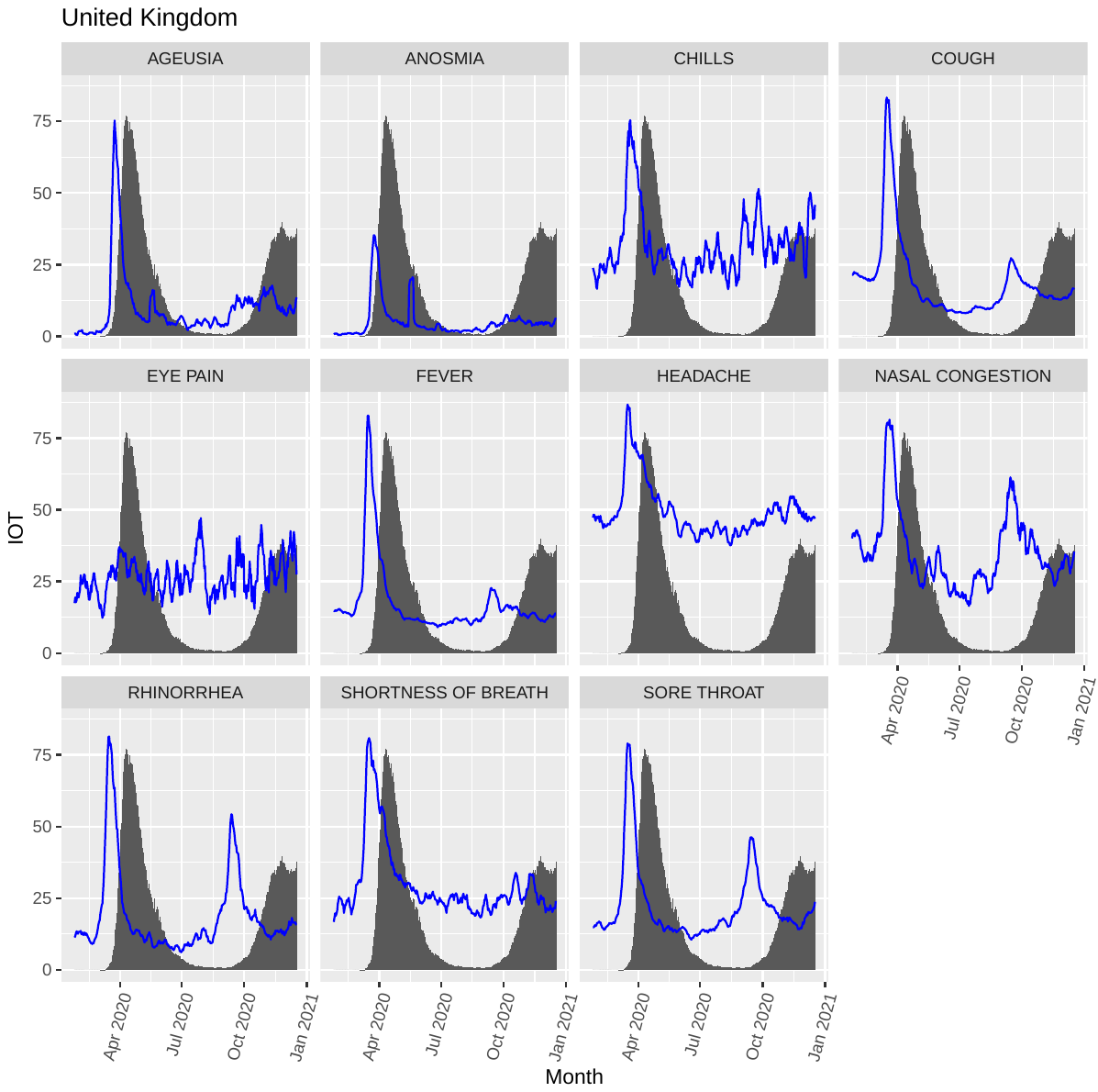

Supplementary Figure 16. Line graphs showing the interest over time (IOT) of the selected topic searches (blue lines) and their relationship with the normalized number of incident deaths per million people (histogram) in the US. Data are plotted as a 7-day moving average to smooth day-by-day fluctuations.

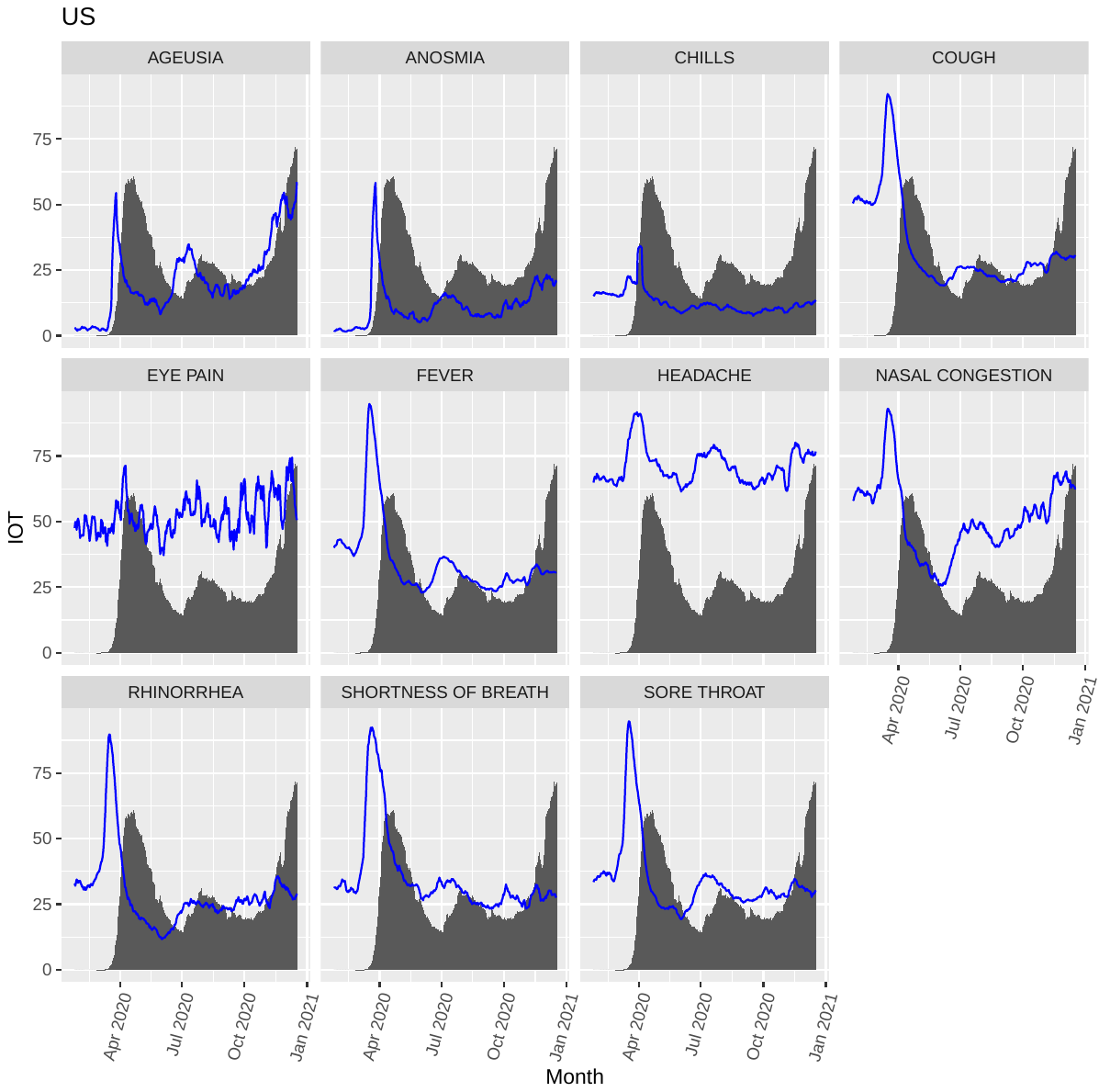

Supplementary Figure 17. Root-mean-square errors (RMSE) of the prediction error for the principal component 2 of the various models for the selected countries. MA1 and MA7 indicate analyses performed on moving average of data 1 day (i.e., original data) and 7 days, respectively. GT indicates models based on both traditional COVID-19 metrics and Google Trends data, while NOGT models based on COVID-19 metrics only. ARIMA: Autoregressive integrated moving average; ETS: Error Trend Seasonality; NNAR: feed-forward neural network autoregression.

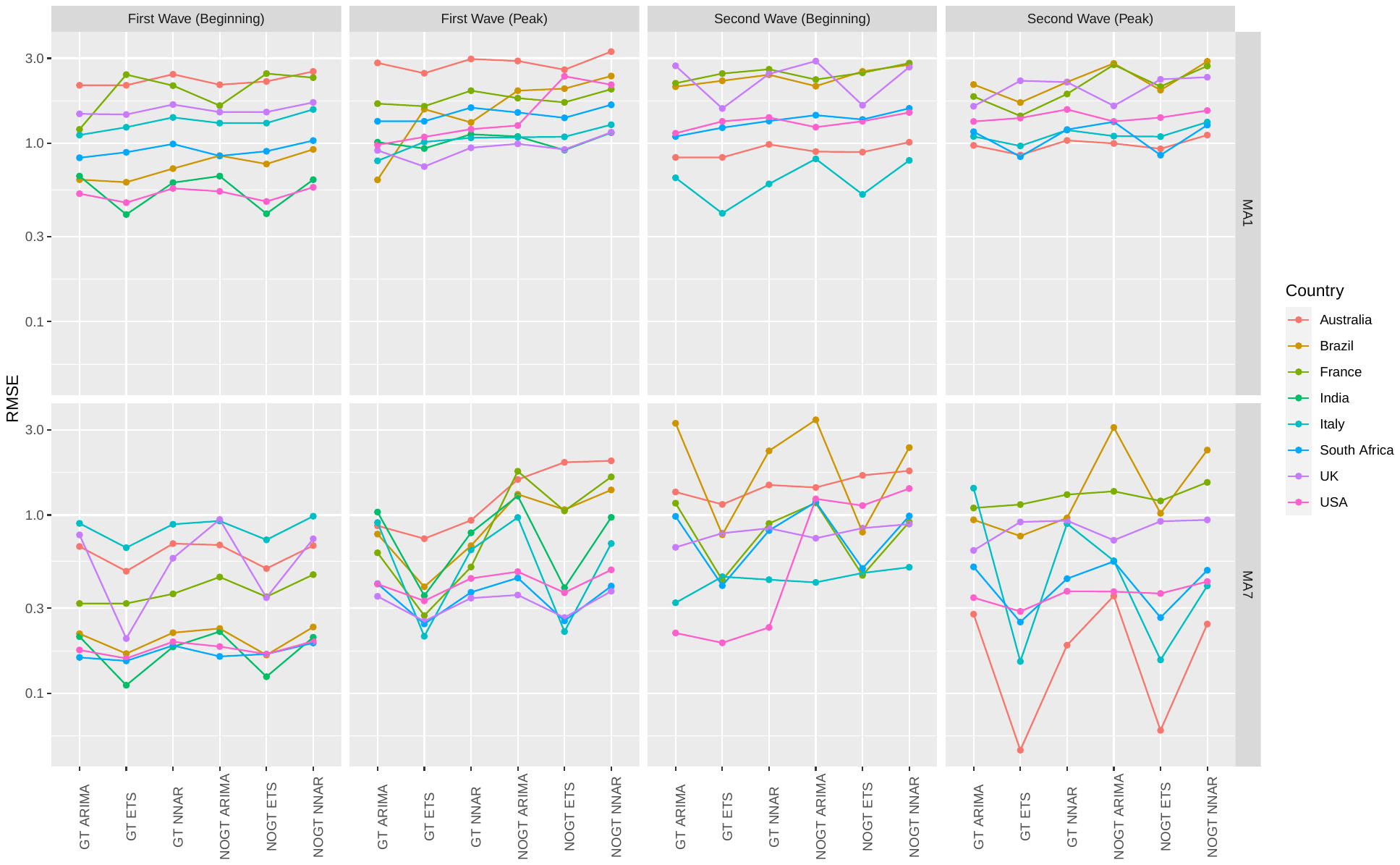

### Supplementary tables

**Supplementary Table 1.** Time-lagged cross-correlations between the 7-day moving average of each topic and the number of new confirmed COVID-19 cases per million population in Australia.

| **LAG** | **Ageusia** | **Anosmia** | **Chills** | **Cough** | **Eye pain** | **Fever** | **Headache** | **Nasal congestion** | **Rhinorrhea** | **Shortness of breath** | **Sore throat** |
| --- | --- | --- | --- | --- | --- | --- | --- | --- | --- | --- | --- |
| 0 | 0.31 | 0.45 | 0.44 | 0.34 | 0.14 | 0.24 | 0.40 | 0.36 | 0.42 | 0.29 | 0.33 |
| -1 | 0.30 | 0.46 | 0.45 | 0.36 | 0.12 | 0.27 | 0.41 | 0.39 | 0.44 | 0.30 | 0.36 |
| -2 | 0.30 | 0.47 | 0.46 | 0.38 | 0.11 | 0.29 | 0.41 | 0.42 | 0.47 | 0.31 | 0.39 |
| -3 | 0.28 | 0.46 | 0.47 | 0.40 | 0.10 | 0.31 | 0.41 | 0.45 | 0.50 | 0.32 | 0.41 |
| -4 | 0.27 | 0.45 | 0.48 | 0.42 | 0.09 | 0.33 | 0.42 | 0.48 | 0.52 | 0.32 | 0.43 |
| -5 | 0.25 | 0.43 | 0.48 | 0.43 | 0.08 | 0.34 | 0.42 | 0.51 | 0.55 | 0.32 | 0.45 |
| -6 | 0.23 | 0.40 | 0.49 | 0.44 | 0.07 | 0.36 | 0.42 | 0.53 | 0.56 | 0.32 | 0.47 |
| -7 | 0.21 | 0.36 | 0.50 | 0.45 | 0.06 | 0.36 | 0.42 | 0.55 | 0.58 | 0.31 | 0.48 |
| -8 | 0.19 | 0.33 | 0.50 | 0.45 | 0.05 | 0.37 | 0.42 | 0.56 | 0.59 | 0.30 | 0.48 |
| -9 | 0.16 | 0.29 | 0.50 | 0.45 | 0.04 | 0.36 | 0.42 | 0.58 | 0.60 | 0.28 | 0.48 |
| -10 | 0.14 | 0.26 | 0.50 | 0.44 | 0.03 | 0.36 | 0.41 | 0.59 | 0.61 | 0.26 | 0.48 |
| -11 | 0.11 | 0.22 | 0.50 | 0.43 | 0.02 | 0.35 | 0.40 | 0.59 | 0.61 | 0.24 | 0.47 |
| -12 | 0.08 | 0.19 | 0.49 | 0.42 | 0.01 | 0.33 | 0.38 | 0.59 | 0.60 | 0.22 | 0.46 |
| -13 | 0.06 | 0.16 | 0.48 | 0.40 | 0.00 | 0.31 | 0.36 | 0.59 | 0.59 | 0.19 | 0.44 |
| -14 | 0.03 | 0.13 | 0.47 | 0.38 | -0.01 | 0.29 | 0.33 | 0.59 | 0.57 | 0.16 | 0.42 |
| -15 | 0.01 | 0.11 | 0.46 | 0.35 | -0.02 | 0.26 | 0.30 | 0.58 | 0.55 | 0.13 | 0.39 |
| -16 | -0.02 | 0.09 | 0.44 | 0.33 | -0.02 | 0.23 | 0.27 | 0.56 | 0.53 | 0.10 | 0.37 |
| -17 | -0.04 | 0.07 | 0.42 | 0.30 | -0.03 | 0.20 | 0.23 | 0.55 | 0.51 | 0.07 | 0.34 |
| -18 | -0.07 | 0.05 | 0.40 | 0.27 | -0.03 | 0.17 | 0.20 | 0.53 | 0.48 | 0.04 | 0.31 |
| -19 | -0.09 | 0.04 | 0.38 | 0.24 | -0.04 | 0.14 | 0.16 | 0.51 | 0.45 | 0.02 | 0.28 |
| -20 | -0.12 | 0.02 | 0.36 | 0.21 | -0.04 | 0.11 | 0.12 | 0.49 | 0.42 | 0.00 | 0.25 |
| -21 | -0.14 | 0.01 | 0.34 | 0.18 | -0.05 | 0.08 | 0.09 | 0.47 | 0.40 | -0.03 | 0.22 |

Data are presented as Pearson's r. Darker shades indicate higher r values.

**Supplementary Table 2.** Time-lagged cross-correlations between the 7-day moving average of each topic and the number of new confirmed COVID-19 cases per million population in Brazil.

| **LAG** | **Ageusia** | **Anosmia** | **Chills** | **Cough** | **Eye pain** | **Fever** | **Headache** | **Nasal congestion** | **Rhinorrhea** | **Shortness of breath** | **Sore throat** |
| --- | --- | --- | --- | --- | --- | --- | --- | --- | --- | --- | --- |
| 0 | 0.35 | 0.03 | -0.47 | -0.54 | -0.44 | -0.5 | -0.02 | -0.27 | -0.16 | -0.44 | -0.29 |
| -1 | 0.36 | 0.04 | -0.46 | -0.54 | -0.44 | -0.49 | -0.01 | -0.26 | -0.15 | -0.44 | -0.29 |
| -2 | 0.37 | 0.05 | -0.46 | -0.53 | -0.44 | -0.48 | 0 | -0.26 | -0.15 | -0.43 | -0.29 |
| -3 | 0.38 | 0.07 | -0.45 | -0.53 | -0.44 | -0.48 | 0.02 | -0.25 | -0.14 | -0.42 | -0.29 |
| -4 | 0.39 | 0.08 | -0.45 | -0.53 | -0.43 | -0.47 | 0.03 | -0.25 | -0.14 | -0.42 | -0.28 |
| -5 | 0.4 | 0.09 | -0.45 | -0.53 | -0.43 | -0.47 | 0.04 | -0.24 | -0.14 | -0.41 | -0.28 |
| -6 | 0.41 | 0.1 | -0.44 | -0.53 | -0.43 | -0.46 | 0.06 | -0.24 | -0.14 | -0.4 | -0.28 |
| -7 | 0.42 | 0.11 | -0.45 | -0.52 | -0.43 | -0.45 | 0.07 | -0.24 | -0.14 | -0.39 | -0.27 |
| -8 | 0.43 | 0.12 | -0.45 | -0.52 | -0.43 | -0.45 | 0.08 | -0.23 | -0.14 | -0.39 | -0.27 |
| -9 | 0.44 | 0.13 | -0.45 | -0.52 | -0.42 | -0.44 | 0.1 | -0.23 | -0.14 | -0.38 | -0.27 |
| -10 | 0.44 | 0.14 | -0.45 | -0.51 | -0.42 | -0.43 | 0.11 | -0.23 | -0.14 | -0.37 | -0.27 |
| -11 | 0.45 | 0.15 | -0.45 | -0.51 | -0.42 | -0.42 | 0.12 | -0.23 | -0.14 | -0.36 | -0.27 |
| -12 | 0.46 | 0.16 | -0.45 | -0.51 | -0.41 | -0.42 | 0.13 | -0.23 | -0.14 | -0.35 | -0.27 |
| -13 | 0.47 | 0.16 | -0.44 | -0.51 | -0.41 | -0.41 | 0.14 | -0.22 | -0.14 | -0.35 | -0.27 |
| -14 | 0.48 | 0.17 | -0.43 | -0.5 | -0.4 | -0.4 | 0.15 | -0.22 | -0.14 | -0.34 | -0.27 |
| -15 | 0.48 | 0.18 | -0.41 | -0.5 | -0.39 | -0.39 | 0.15 | -0.21 | -0.13 | -0.33 | -0.27 |
| -16 | 0.49 | 0.19 | -0.4 | -0.5 | -0.38 | -0.38 | 0.16 | -0.21 | -0.11 | -0.32 | -0.27 |
| -17 | 0.49 | 0.2 | -0.38 | -0.5 | -0.36 | -0.38 | 0.16 | -0.21 | -0.1 | -0.31 | -0.27 |
| -18 | 0.5 | 0.21 | -0.37 | -0.49 | -0.35 | -0.37 | 0.17 | -0.21 | -0.09 | -0.3 | -0.27 |
| -19 | 0.5 | 0.22 | -0.35 | -0.49 | -0.34 | -0.37 | 0.17 | -0.21 | -0.08 | -0.29 | -0.27 |
| -20 | 0.5 | 0.23 | -0.33 | -0.49 | -0.32 | -0.36 | 0.18 | -0.21 | -0.08 | -0.28 | -0.27 |
| -21 | 0.5 | 0.24 | -0.32 | -0.48 | -0.31 | -0.36 | 0.18 | -0.21 | -0.07 | -0.28 | -0.27 |

Data are presented as Pearson's r. Darker shades indicate higher r values.

**Supplementary Table 3.** Time-lagged cross-correlations between the 7-day moving average of each topic and the number of new confirmed COVID-19 cases per million population in France.

| **LAG** | **Ageusia** | **Anosmia** | **Chills** | **Cough** | **Eye pain** | **Fever** | **Headache** | **Nasal congestion** | **Rhinorrhea** | **Shortness of breath** | **Sore throat** |
| --- | --- | --- | --- | --- | --- | --- | --- | --- | --- | --- | --- |
| 0 | 0.53 | 0.23 | -0.19 | -0.1 | -0.15 | -0.14 | 0.28 | 0.21 | -0.05 | -0.16 | 0.01 |
| -1 | 0.53 | 0.24 | -0.2 | -0.1 | -0.15 | -0.14 | 0.28 | 0.22 | -0.04 | -0.16 | 0.03 |
| -2 | 0.54 | 0.24 | -0.2 | -0.09 | -0.15 | -0.13 | 0.27 | 0.24 | -0.03 | -0.15 | 0.04 |
| -3 | 0.55 | 0.25 | -0.2 | -0.09 | -0.15 | -0.12 | 0.27 | 0.26 | -0.01 | -0.15 | 0.06 |
| -4 | 0.55 | 0.25 | -0.21 | -0.08 | -0.15 | -0.12 | 0.27 | 0.27 | 0 | -0.15 | 0.07 |
| -5 | 0.55 | 0.26 | -0.22 | -0.08 | -0.16 | -0.11 | 0.27 | 0.29 | 0.01 | -0.15 | 0.08 |
| -6 | 0.55 | 0.26 | -0.22 | -0.07 | -0.17 | -0.1 | 0.26 | 0.31 | 0.03 | -0.15 | 0.1 |
| -7 | 0.55 | 0.26 | -0.22 | -0.07 | -0.17 | -0.1 | 0.26 | 0.33 | 0.04 | -0.15 | 0.11 |
| -8 | 0.54 | 0.26 | -0.22 | -0.06 | -0.16 | -0.09 | 0.25 | 0.34 | 0.05 | -0.15 | 0.12 |
| -9 | 0.54 | 0.25 | -0.23 | -0.06 | -0.16 | -0.09 | 0.25 | 0.35 | 0.07 | -0.15 | 0.13 |
| -10 | 0.53 | 0.25 | -0.23 | -0.05 | -0.17 | -0.08 | 0.24 | 0.37 | 0.08 | -0.14 | 0.15 |
| -11 | 0.51 | 0.24 | -0.23 | -0.05 | -0.18 | -0.08 | 0.23 | 0.38 | 0.09 | -0.14 | 0.16 |
| -12 | 0.5 | 0.23 | -0.23 | -0.04 | -0.18 | -0.07 | 0.23 | 0.39 | 0.1 | -0.13 | 0.17 |
| -13 | 0.49 | 0.23 | -0.23 | -0.04 | -0.18 | -0.07 | 0.22 | 0.39 | 0.1 | -0.13 | 0.18 |
| -14 | 0.49 | 0.23 | -0.23 | -0.04 | -0.18 | -0.06 | 0.21 | 0.4 | 0.11 | -0.13 | 0.18 |
| -15 | 0.48 | 0.23 | -0.23 | -0.04 | -0.2 | -0.06 | 0.2 | 0.4 | 0.12 | -0.13 | 0.19 |
| -16 | 0.48 | 0.24 | -0.24 | -0.04 | -0.21 | -0.06 | 0.19 | 0.41 | 0.12 | -0.13 | 0.2 |
| -17 | 0.48 | 0.25 | -0.25 | -0.04 | -0.21 | -0.06 | 0.19 | 0.41 | 0.13 | -0.13 | 0.21 |
| -18 | 0.48 | 0.26 | -0.26 | -0.04 | -0.21 | -0.06 | 0.18 | 0.42 | 0.13 | -0.14 | 0.22 |
| -19 | 0.48 | 0.27 | -0.26 | -0.04 | -0.22 | -0.06 | 0.17 | 0.42 | 0.14 | -0.14 | 0.22 |
| -20 | 0.47 | 0.28 | -0.27 | -0.04 | -0.23 | -0.06 | 0.16 | 0.42 | 0.15 | -0.15 | 0.23 |
| -21 | 0.46 | 0.29 | -0.27 | -0.04 | -0.23 | -0.05 | 0.15 | 0.43 | 0.16 | -0.15 | 0.24 |

Data are presented as Pearson's r. Darker shades indicate higher r values.

**Supplementary Table 4.** Time-lagged cross-correlations between the 7-day moving average of each topic and the number of new confirmed COVID-19 cases per million population in India.

| **LAG** | **Ageusia** | **Anosmia** | **Chills** | **Cough** | **Eye pain** | **Fever** | **Headache** | **Nasal congestion** | **Rhinorrhea** | **Shortness of breath** | **Sore throat** |
| --- | --- | --- | --- | --- | --- | --- | --- | --- | --- | --- | --- |
| 0 | 0.89 | 0.91 | 0.19 | -0.37 | 0.47 | 0.52 | 0.23 | -0.48 | -0.12 | -0.22 | -0.29 |
| -1 | 0.9 | 0.92 | 0.2 | -0.38 | 0.48 | 0.53 | 0.25 | -0.47 | -0.12 | -0.21 | -0.29 |
| -2 | 0.9 | 0.92 | 0.2 | -0.38 | 0.49 | 0.54 | 0.26 | -0.47 | -0.12 | -0.2 | -0.28 |
| -3 | 0.9 | 0.92 | 0.21 | -0.38 | 0.49 | 0.55 | 0.27 | -0.47 | -0.13 | -0.19 | -0.28 |
| -4 | 0.9 | 0.93 | 0.21 | -0.38 | 0.5 | 0.55 | 0.28 | -0.47 | -0.13 | -0.18 | -0.27 |
| -5 | 0.91 | 0.93 | 0.22 | -0.39 | 0.5 | 0.56 | 0.3 | -0.47 | -0.13 | -0.18 | -0.27 |
| -6 | 0.91 | 0.93 | 0.22 | -0.39 | 0.51 | 0.57 | 0.31 | -0.47 | -0.13 | -0.17 | -0.26 |
| -7 | 0.91 | 0.93 | 0.22 | -0.39 | 0.52 | 0.57 | 0.32 | -0.47 | -0.13 | -0.16 | -0.26 |
| -8 | 0.91 | 0.93 | 0.23 | -0.39 | 0.52 | 0.57 | 0.33 | -0.47 | -0.13 | -0.15 | -0.25 |
| -9 | 0.91 | 0.93 | 0.23 | -0.4 | 0.53 | 0.58 | 0.34 | -0.47 | -0.14 | -0.14 | -0.25 |
| -10 | 0.91 | 0.93 | 0.23 | -0.4 | 0.54 | 0.58 | 0.34 | -0.47 | -0.14 | -0.13 | -0.24 |
| -11 | 0.91 | 0.92 | 0.24 | -0.41 | 0.55 | 0.58 | 0.35 | -0.47 | -0.14 | -0.12 | -0.24 |
| -12 | 0.9 | 0.92 | 0.24 | -0.41 | 0.55 | 0.59 | 0.36 | -0.47 | -0.14 | -0.11 | -0.23 |
| -13 | 0.9 | 0.92 | 0.24 | -0.41 | 0.56 | 0.59 | 0.37 | -0.47 | -0.14 | -0.11 | -0.23 |
| -14 | 0.9 | 0.91 | 0.25 | -0.42 | 0.57 | 0.59 | 0.38 | -0.47 | -0.14 | -0.1 | -0.23 |
| -15 | 0.9 | 0.91 | 0.25 | -0.42 | 0.57 | 0.59 | 0.38 | -0.47 | -0.14 | -0.09 | -0.22 |
| -16 | 0.89 | 0.9 | 0.25 | -0.43 | 0.58 | 0.59 | 0.39 | -0.47 | -0.15 | -0.08 | -0.22 |
| -17 | 0.89 | 0.9 | 0.25 | -0.43 | 0.58 | 0.59 | 0.39 | -0.46 | -0.15 | -0.07 | -0.21 |
| -18 | 0.89 | 0.89 | 0.26 | -0.44 | 0.58 | 0.59 | 0.4 | -0.46 | -0.15 | -0.06 | -0.21 |
| -19 | 0.88 | 0.89 | 0.26 | -0.44 | 0.59 | 0.59 | 0.4 | -0.46 | -0.15 | -0.05 | -0.21 |
| -20 | 0.88 | 0.88 | 0.26 | -0.45 | 0.59 | 0.59 | 0.41 | -0.46 | -0.16 | -0.04 | -0.2 |
| -21 | 0.88 | 0.88 | 0.27 | -0.45 | 0.59 | 0.59 | 0.41 | -0.46 | -0.16 | -0.04 | -0.2 |

Data are presented as Pearson's r. Darker shades indicate higher r values.

**Supplementary Table 5.** Time-lagged cross-correlations between the 7-day moving average of each topic and the number of new confirmed COVID-19 cases per million population in Italy.

| **LAG** | **Ageusia** | **Anosmia** | **Chills** | **Cough** | **Eye pain** | **Fever** | **Headache** | **Nasal congestion** | **Rhinorrhea** | **Shortness of breath** | **Sore throat** |
| --- | --- | --- | --- | --- | --- | --- | --- | --- | --- | --- | --- |
| 0 | 0.69 | 0.49 | 0.09 | 0.08 | 0.07 | 0.17 | 0.81 | 0.24 | 0 | -0.02 | 0.22 |
| -1 | 0.71 | 0.51 | 0.09 | 0.09 | 0.08 | 0.19 | 0.82 | 0.24 | 0.01 | -0.01 | 0.23 |
| -2 | 0.72 | 0.53 | 0.1 | 0.1 | 0.08 | 0.21 | 0.84 | 0.25 | 0.01 | 0 | 0.24 |
| -3 | 0.74 | 0.54 | 0.1 | 0.12 | 0.09 | 0.23 | 0.85 | 0.25 | 0.01 | 0 | 0.25 |
| -4 | 0.76 | 0.56 | 0.1 | 0.13 | 0.1 | 0.24 | 0.86 | 0.25 | 0.02 | 0.01 | 0.26 |
| -5 | 0.77 | 0.57 | 0.11 | 0.14 | 0.1 | 0.26 | 0.87 | 0.25 | 0.02 | 0.01 | 0.27 |
| -6 | 0.78 | 0.59 | 0.11 | 0.15 | 0.11 | 0.27 | 0.87 | 0.25 | 0.02 | 0.02 | 0.28 |
| -7 | 0.79 | 0.6 | 0.11 | 0.16 | 0.11 | 0.29 | 0.87 | 0.26 | 0.02 | 0.02 | 0.29 |
| -8 | 0.8 | 0.61 | 0.11 | 0.17 | 0.11 | 0.3 | 0.87 | 0.26 | 0.02 | 0.03 | 0.3 |
| -9 | 0.8 | 0.61 | 0.11 | 0.17 | 0.11 | 0.31 | 0.87 | 0.26 | 0.02 | 0.03 | 0.31 |
| -10 | 0.81 | 0.62 | 0.11 | 0.18 | 0.12 | 0.32 | 0.86 | 0.26 | 0.02 | 0.03 | 0.32 |
| -11 | 0.81 | 0.62 | 0.11 | 0.19 | 0.12 | 0.33 | 0.86 | 0.26 | 0.02 | 0.03 | 0.33 |
| -12 | 0.81 | 0.62 | 0.11 | 0.19 | 0.12 | 0.34 | 0.85 | 0.26 | 0.02 | 0.03 | 0.34 |
| -13 | 0.81 | 0.62 | 0.11 | 0.2 | 0.13 | 0.34 | 0.85 | 0.26 | 0.02 | 0.03 | 0.36 |
| -14 | 0.8 | 0.62 | 0.11 | 0.2 | 0.13 | 0.35 | 0.84 | 0.25 | 0.02 | 0.03 | 0.37 |
| -15 | 0.8 | 0.62 | 0.1 | 0.21 | 0.13 | 0.35 | 0.83 | 0.25 | 0.02 | 0.03 | 0.38 |
| -16 | 0.79 | 0.62 | 0.1 | 0.21 | 0.13 | 0.36 | 0.82 | 0.24 | 0.02 | 0.03 | 0.4 |
| -17 | 0.78 | 0.61 | 0.09 | 0.21 | 0.13 | 0.36 | 0.8 | 0.23 | 0.02 | 0.02 | 0.41 |
| -18 | 0.77 | 0.6 | 0.09 | 0.21 | 0.12 | 0.36 | 0.79 | 0.23 | 0.02 | 0.02 | 0.42 |
| -19 | 0.75 | 0.6 | 0.08 | 0.21 | 0.12 | 0.36 | 0.77 | 0.22 | 0.02 | 0.01 | 0.44 |
| -20 | 0.74 | 0.58 | 0.08 | 0.21 | 0.11 | 0.36 | 0.75 | 0.22 | 0.02 | 0.01 | 0.45 |
| -21 | 0.72 | 0.57 | 0.07 | 0.21 | 0.11 | 0.36 | 0.73 | 0.21 | 0.02 | 0 | 0.46 |

Data are presented as Pearson's r. Darker shades indicate higher r values.

**Supplementary Table 6.** Time-lagged cross-correlations between the 7-day moving average of each topic and the number of new confirmed COVID-19 cases per million population in South Africa.

| **LAG** | **Ageusia** | **Anosmia** | **Chills** | **Cough** | **Eye pain** | **Fever** | **Headache** | **Nasal congestion** | **Rhinorrhea** | **Shortness of breath** | **Sore throat** |
| --- | --- | --- | --- | --- | --- | --- | --- | --- | --- | --- | --- |
| 0 | 0.91 | 0.92 | 0.45 | 0.16 | 0.26 | 0.02 | 0.34 | 0.45 | 0.3 | 0.19 | -0.15 |
| -1 | 0.91 | 0.91 | 0.46 | 0.17 | 0.27 | 0.02 | 0.35 | 0.46 | 0.3 | 0.19 | -0.14 |
| -2 | 0.9 | 0.91 | 0.48 | 0.17 | 0.29 | 0.02 | 0.35 | 0.46 | 0.3 | 0.19 | -0.13 |
| -3 | 0.9 | 0.91 | 0.49 | 0.17 | 0.3 | 0.03 | 0.34 | 0.47 | 0.29 | 0.19 | -0.13 |
| -4 | 0.89 | 0.91 | 0.5 | 0.17 | 0.31 | 0.03 | 0.34 | 0.48 | 0.29 | 0.18 | -0.12 |
| -5 | 0.88 | 0.9 | 0.51 | 0.17 | 0.32 | 0.03 | 0.33 | 0.48 | 0.28 | 0.18 | -0.11 |
| -6 | 0.88 | 0.9 | 0.52 | 0.17 | 0.33 | 0.03 | 0.32 | 0.49 | 0.28 | 0.18 | -0.11 |
| -7 | 0.87 | 0.89 | 0.53 | 0.17 | 0.34 | 0.03 | 0.31 | 0.49 | 0.27 | 0.17 | -0.11 |
| -8 | 0.86 | 0.88 | 0.54 | 0.16 | 0.35 | 0.03 | 0.3 | 0.49 | 0.27 | 0.16 | -0.1 |
| -9 | 0.85 | 0.87 | 0.55 | 0.16 | 0.35 | 0.03 | 0.29 | 0.49 | 0.26 | 0.16 | -0.1 |
| -10 | 0.84 | 0.86 | 0.55 | 0.16 | 0.35 | 0.03 | 0.29 | 0.48 | 0.25 | 0.14 | -0.1 |
| -11 | 0.83 | 0.84 | 0.56 | 0.15 | 0.35 | 0.03 | 0.28 | 0.48 | 0.23 | 0.13 | -0.1 |
| -12 | 0.81 | 0.83 | 0.56 | 0.15 | 0.34 | 0.03 | 0.27 | 0.47 | 0.22 | 0.13 | -0.1 |
| -13 | 0.8 | 0.81 | 0.56 | 0.14 | 0.34 | 0.03 | 0.26 | 0.46 | 0.21 | 0.12 | -0.1 |
| -14 | 0.78 | 0.79 | 0.56 | 0.13 | 0.33 | 0.03 | 0.26 | 0.45 | 0.19 | 0.11 | -0.11 |
| -15 | 0.76 | 0.77 | 0.55 | 0.12 | 0.32 | 0.02 | 0.25 | 0.44 | 0.18 | 0.1 | -0.11 |
| -16 | 0.74 | 0.75 | 0.55 | 0.11 | 0.31 | 0.02 | 0.25 | 0.43 | 0.16 | 0.09 | -0.11 |
| -17 | 0.72 | 0.73 | 0.55 | 0.1 | 0.29 | 0.02 | 0.25 | 0.41 | 0.15 | 0.08 | -0.11 |
| -18 | 0.7 | 0.7 | 0.54 | 0.09 | 0.27 | 0.02 | 0.24 | 0.4 | 0.14 | 0.08 | -0.12 |
| -19 | 0.67 | 0.68 | 0.54 | 0.08 | 0.25 | 0.02 | 0.24 | 0.39 | 0.12 | 0.07 | -0.12 |
| -20 | 0.65 | 0.66 | 0.53 | 0.07 | 0.24 | 0.01 | 0.24 | 0.38 | 0.11 | 0.05 | -0.12 |
| -21 | 0.62 | 0.64 | 0.52 | 0.06 | 0.23 | 0.01 | 0.23 | 0.36 | 0.1 | 0.04 | -0.13 |

Data are presented as Pearson's r. Darker shades indicate higher r values.

**Supplementary Table 7.** Time-lagged cross-correlations between the 7-day moving average of each topic and the number of new confirmed COVID-19 cases per million population in the UK.

| **LAG** | **Ageusia** | **Anosmia** | **Chills** | **Cough** | **Eye pain** | **Fever** | **Headache** | **Nasal congestion** | **Rhinorrhea** | **Shortness of breath** | **Sore throat** |
| --- | --- | --- | --- | --- | --- | --- | --- | --- | --- | --- | --- |
| 0 | 0.17 | 0.07 | 0.15 | -0.13 | 0.32 | -0.13 | 0.07 | -0.07 | -0.09 | -0.09 | -0.09 |
| -1 | 0.18 | 0.07 | 0.15 | -0.12 | 0.33 | -0.13 | 0.07 | -0.06 | -0.08 | -0.08 | -0.08 |
| -2 | 0.18 | 0.07 | 0.15 | -0.11 | 0.33 | -0.12 | 0.07 | -0.05 | -0.07 | -0.08 | -0.07 |
| -3 | 0.18 | 0.08 | 0.16 | -0.11 | 0.33 | -0.11 | 0.08 | -0.04 | -0.06 | -0.07 | -0.06 |
| -4 | 0.19 | 0.08 | 0.17 | -0.1 | 0.32 | -0.11 | 0.08 | -0.02 | -0.05 | -0.07 | -0.05 |
| -5 | 0.19 | 0.08 | 0.17 | -0.09 | 0.3 | -0.1 | 0.08 | -0.01 | -0.04 | -0.06 | -0.03 |
| -6 | 0.2 | 0.09 | 0.18 | -0.08 | 0.3 | -0.09 | 0.08 | 0.01 | -0.02 | -0.06 | -0.02 |
| -7 | 0.2 | 0.09 | 0.18 | -0.07 | 0.3 | -0.08 | 0.08 | 0.02 | -0.01 | -0.05 | -0.01 |
| -8 | 0.2 | 0.09 | 0.18 | -0.06 | 0.29 | -0.07 | 0.08 | 0.04 | 0 | -0.05 | 0 |
| -9 | 0.21 | 0.1 | 0.18 | -0.06 | 0.28 | -0.07 | 0.08 | 0.05 | 0.01 | -0.05 | 0.01 |
| -10 | 0.21 | 0.1 | 0.17 | -0.05 | 0.27 | -0.06 | 0.08 | 0.07 | 0.03 | -0.05 | 0.03 |
| -11 | 0.2 | 0.1 | 0.17 | -0.04 | 0.26 | -0.05 | 0.08 | 0.08 | 0.04 | -0.04 | 0.04 |
| -12 | 0.2 | 0.1 | 0.17 | -0.03 | 0.25 | -0.04 | 0.08 | 0.09 | 0.05 | -0.04 | 0.05 |
| -13 | 0.2 | 0.09 | 0.18 | -0.02 | 0.24 | -0.04 | 0.08 | 0.1 | 0.07 | -0.04 | 0.06 |
| -14 | 0.2 | 0.09 | 0.19 | -0.01 | 0.24 | -0.03 | 0.08 | 0.11 | 0.08 | -0.04 | 0.07 |
| -15 | 0.2 | 0.09 | 0.2 | -0.01 | 0.24 | -0.02 | 0.08 | 0.12 | 0.1 | -0.04 | 0.09 |
| -16 | 0.2 | 0.09 | 0.21 | 0 | 0.24 | -0.02 | 0.08 | 0.14 | 0.11 | -0.04 | 0.1 |
| -17 | 0.19 | 0.09 | 0.22 | 0.01 | 0.24 | -0.01 | 0.08 | 0.15 | 0.13 | -0.04 | 0.11 |
| -18 | 0.19 | 0.09 | 0.23 | 0.01 | 0.24 | 0 | 0.08 | 0.16 | 0.14 | -0.04 | 0.12 |
| -19 | 0.19 | 0.08 | 0.23 | 0.02 | 0.24 | 0 | 0.08 | 0.17 | 0.15 | -0.04 | 0.13 |
| -20 | 0.19 | 0.08 | 0.23 | 0.02 | 0.23 | 0.01 | 0.08 | 0.18 | 0.17 | -0.04 | 0.14 |
| -21 | 0.18 | 0.08 | 0.22 | 0.03 | 0.21 | 0.01 | 0.07 | 0.19 | 0.18 | -0.04 | 0.15 |

Data are presented as Pearson's r. Darker shades indicate higher r values.

**Supplementary Table 8.** Time-lagged cross-correlations between the 7-day moving average of each topic and the number of new confirmed COVID-19 cases per million population in the US.

| **LAG** | **Ageusia** | **Anosmia** | **Chills** | **Cough** | **Eye pain** | **Fever** | **Headache** | **Nasal congestion** | **Rhinorrhea** | **Shortness of breath** | **Sore throat** |
| --- | --- | --- | --- | --- | --- | --- | --- | --- | --- | --- | --- |
| 0 | 0.84 | 0.43 | -0.26 | -0.32 | 0.57 | -0.27 | 0.25 | 0.21 | -0.12 | -0.29 | -0.22 |
| -1 | 0.83 | 0.43 | -0.26 | -0.32 | 0.58 | -0.27 | 0.25 | 0.21 | -0.11 | -0.28 | -0.21 |
| -2 | 0.82 | 0.43 | -0.26 | -0.31 | 0.59 | -0.27 | 0.25 | 0.21 | -0.11 | -0.28 | -0.21 |
| -3 | 0.81 | 0.44 | -0.26 | -0.31 | 0.59 | -0.26 | 0.24 | 0.21 | -0.1 | -0.28 | -0.2 |
| -4 | 0.81 | 0.44 | -0.27 | -0.31 | 0.59 | -0.26 | 0.24 | 0.21 | -0.1 | -0.27 | -0.2 |
| -5 | 0.8 | 0.44 | -0.27 | -0.3 | 0.58 | -0.25 | 0.23 | 0.21 | -0.09 | -0.27 | -0.19 |
| -6 | 0.79 | 0.43 | -0.27 | -0.3 | 0.57 | -0.25 | 0.23 | 0.2 | -0.09 | -0.27 | -0.18 |
| -7 | 0.79 | 0.43 | -0.27 | -0.3 | 0.55 | -0.25 | 0.22 | 0.2 | -0.08 | -0.26 | -0.18 |
| -8 | 0.78 | 0.42 | -0.27 | -0.3 | 0.52 | -0.24 | 0.21 | 0.2 | -0.08 | -0.26 | -0.17 |
| -9 | 0.77 | 0.42 | -0.28 | -0.3 | 0.51 | -0.24 | 0.2 | 0.2 | -0.08 | -0.26 | -0.17 |
| -10 | 0.76 | 0.41 | -0.28 | -0.29 | 0.49 | -0.24 | 0.19 | 0.19 | -0.07 | -0.26 | -0.17 |
| -11 | 0.75 | 0.4 | -0.29 | -0.29 | 0.47 | -0.23 | 0.19 | 0.19 | -0.07 | -0.26 | -0.16 |
| -12 | 0.74 | 0.39 | -0.29 | -0.29 | 0.45 | -0.23 | 0.17 | 0.18 | -0.06 | -0.25 | -0.16 |
| -13 | 0.73 | 0.38 | -0.29 | -0.29 | 0.43 | -0.23 | 0.17 | 0.18 | -0.06 | -0.25 | -0.16 |
| -14 | 0.71 | 0.37 | -0.3 | -0.28 | 0.41 | -0.23 | 0.16 | 0.17 | -0.06 | -0.25 | -0.15 |
| -15 | 0.7 | 0.36 | -0.3 | -0.28 | 0.4 | -0.23 | 0.15 | 0.17 | -0.06 | -0.25 | -0.15 |
| -16 | 0.68 | 0.35 | -0.3 | -0.28 | 0.38 | -0.23 | 0.14 | 0.16 | -0.06 | -0.25 | -0.15 |
| -17 | 0.66 | 0.33 | -0.31 | -0.28 | 0.37 | -0.23 | 0.13 | 0.15 | -0.06 | -0.25 | -0.15 |
| -18 | 0.64 | 0.32 | -0.31 | -0.28 | 0.36 | -0.23 | 0.12 | 0.15 | -0.06 | -0.25 | -0.15 |
| -19 | 0.62 | 0.31 | -0.31 | -0.28 | 0.37 | -0.23 | 0.11 | 0.14 | -0.06 | -0.25 | -0.15 |
| -20 | 0.6 | 0.3 | -0.31 | -0.28 | 0.37 | -0.23 | 0.11 | 0.13 | -0.06 | -0.25 | -0.15 |
| -21 | 0.58 | 0.29 | -0.32 | -0.28 | 0.38 | -0.23 | 0.1 | 0.12 | -0.07 | -0.25 | -0.15 |

Data are presented as Pearson's r. Darker shades indicate higher r values.
